# Aperiodic subthalamic activity underlies sleep-wake modulation of beta power during conventional and adaptive deep brain stimulation in Parkinson’s disease

**DOI:** 10.64898/2026.05.11.26352836

**Authors:** Laura Caffi, Fabrizio Luiso, Simona Cascino, Rita Habib, Salvatore Bonvegna, Pasquale Serrao, Emma Crespi, Sara Marceglia, Chiara Palmisano, Alberto Mazzoni, Ioannis U. Isaias

## Abstract

Subthalamic local field potential (STN-LFP) activity within patient-specific beta frequency ranges is an established biomarker for adaptive deep brain stimulation (aDBS) in Parkinson’s disease (PD). While beta power correlates with akinetic-rigid symptoms, it is also modulated by physiological states such as sleep, highlighting the importance of understanding state-dependent spectral dynamics for adaptive stimulation. We continuously recorded STN-LFP spectra in ten PD patients over two weeks in both conventional and adaptive DBS. We show that sleep-related reductions in beta power occur independently of stimulation mode and are primarily associated with broadband aperiodic spectral changes across both low and high beta bands rather than periodic beta oscillations. These findings support the integration of aperiodic spectral features into future adaptive neuromodulation algorithms to improve biomarker specificity and optimize aDBS in PD.

## INTRODUCTION

Deep brain stimulation (DBS) of the subthalamic nucleus (STN) is a well-established treatment for patients with Parkinson’s disease (PD) [1]. The recent introduction of new implantable pulse generators (IPG) capable of continuously recording local field potentials (LFP) from chronically stimulated brain nuclei has enabled the collection of critical data to advance our understanding of PD pathophysiology [2]. This technological development has paved the way for novel stimulation paradigms, such as adaptive DBS (aDBS), which uses patient-specific real-time LFP to automatically adjust stimulation amplitude [3–5]. To date, the most widely studied biomarker for aDBS in PD is subthalamic oscillatory activity within the beta frequency band (12–30 Hz) [6]. Beta power within a patient-specific frequency range has been associated with the severity of akinetic-rigid symptoms [7], as well as with clinical improvement following levodopa administration [8] and STN-DBS [9]. The presence of a prominent subthalamic beta peak across contacts can further guide the selection of the optimal stimulation contact [10].

Sleep represents a particularly compelling area for further investigation, not only due to its significant clinical relevance in PD [11,12], but also because of its potential as a novel target for adaptive neuromodulation. Modulation of STN activity across the sleep-wake cycle has been documented both in clinical settings using externalized electrodes, and chronically in home settings prior to DBS activation or with optimized cDBS. These studies consistently demonstrate reduced beta activity in the STN during non-rapid-eye-movement (NREM) sleep compared with wakefulness, whereas beta power during rapid-eye-movement (REM) sleep is similar to, or occasionally higher, that observed during wakefulness [13–20]. Patients with PD show disturbances across nearly all aspects of sleep, including sleep architecture and parasomnias during both REM and NREM sleep [21–23]. The most common symptoms are delayed sleep onset and increased wakefulness after sleep onset (WASO), resulting in reduced total sleep time and excessive daytime sleepiness [23–25]. Moreover, total and percentage time spent in deep NREM (stages N2 and N3) and REM sleep is reduced in PD [26–29]. Importantly, REM behaviour disorder (RBD) has been recognized as a prodromal symptom of PD [30,31]. Deep sleep has been associated with the clearance of metabolic waste products, including alpha-synuclein [32,33], maintenance of synaptic homeostasis and memory consolidation [34–40]. Therefore, reduction in deep sleep and increased sleep fragmentation in PD may contribute to worsening symptoms, accelerate disease progression [41], and impair cognition, potentially leading to future cognitive decline or dementia. Consequently, sleep disturbances have a substantial impact on the quality of life of both patients and caregivers, making sleep a crucial therapeutic target [21,42,43].

Conventional DBS (cDBS), based on fixed stimulation parameters over time, has been shown to improve sleep quality and efficiency by increasing total sleep time and decreasing sleep latency and WASO [44]. The REM phase appears to be improved consistently, with reduced REM latency and increases in both REM duration and percentage [45]. However, DBS does not seem to improve RBD [44,46], and findings regarding improvements in NREM sleep remain inconsistent across studies [45]. Currently, cDBS parameters are primarily optimized to control daytime motor symptoms. Tailoring stimulation settings during sleep, or dynamically adapting stimulation according to specific sleep stages, may further enhance the therapeutic efficacy of DBS and improve nocturnal symptom management. Existing aDBS systems [3–5,47–49] rely on algorithms designed to detect slow neural signal fluctuations and sustained state transitions, including changes between “ON” and “OFF” motor states or the emergence of dyskinesias. Within this framework, sleep represents a particularly relevant physiological context, as its prolonged and well-defined stages may provide a suitable substrate for the implementation and optimization of currently available adaptive neuromodulation technologies, including for example, the automatic reduction of stimulation amplitude during the night to prevent overstimulation and associated side effects [50], while also potentially mitigating long-term habituation to therapy for symptoms such as tremor [51].

However, the effects of different stimulation modalities, such as cDBS versus aDBS, on STN activity across the sleep-wake cycle remain unclear and are critical to inform novel aDBS programming paradigms. Moreover, previous studies examined power modulations within conventional frequency bands without distinguishing true oscillatory activity from the underlying aperiodic background. STN power spectra typically exhibit an aperiodic background component characterized by a 1/f^n^ profile, in which power decreases exponentially with increasing frequencies. This component can be described by three parameters: the offset, reflecting broadband power shifts; the slope, corresponding to the exponent of the aperiodic line in log-log space; and the knee which represents the point at which the spectrum bends [52]. These parameters have been linked to distinct physiological mechanisms: the offset is thought to reflect population spiking activity [53], whereas the slope is associated with the balance between excitatory and inhibitory synaptic inputs [54]. Notably, both offset and aperiodic broadband power have been shown to correlate positively with PD motor symptom severity, highlighting their potential as biomarkers for aDBS in PD [55]. In the context of sleep physiology the cortical aperiodic slope has been proposed as a marker of arousal, with steeper slopes observed in deeper sleep states [56–60]. However, subcortical aperiodic activity has not been yet explored in PD during the sleep-wake cycle, warranting further investigation.

In this study, we present continuous recordings of STN activity during both sleep and wake states in eleven patients with PD. Following DBS activation, patients underwent two weeks of cDBS and two weeks of aDBS in a randomized, double-blind, crossover design. We examined differences between sleep and wake states in both oscillatory and aperiodic activity, as well as the effects of cDBS and aDBS. Oscillatory activity was analyzed within canonical frequency bands (low beta, 12–20 Hz; high beta, 21–30 Hz) and within patient-specific frequency ranges centered around the most prominent spectral peaks. We found that patient-specific peaks remained stable across stimulation modes (cDBS/aDBS) and circadian states (sleep/wake). STN activity was higher in the wake compared to the sleep state, primarily driven by differences in aperiodic parameters, with no impact of stimulation mode. These findings support the inclusion of aperiodic spectral components to enhance sensitivity and inform the optimization of future aDBS algorithms.

## METHODS

### Participants and experimental protocol

We enrolled eleven patients with idiopathic PD who received STN-DBS with the AlphaDBS IPG (Newronika S.p.A.) for clinical indications, either at first implantation (de novo) or at battery replacement. Patients participated in a randomized, double-blind crossover trial consisting of two consecutive two-week periods of cDBS and aDBS, while dopaminergic medications were kept unchanged [61]. De novo patients entered the study after an approximately five-week postoperative healing period during which stimulation was kept off.

This study extends the findings of a previous clinical trial (NCT04681534) [5], which demonstrated the safety and efficacy of chronic aDBS delivered through a linear proportional algorithm using the AlphaDBS device. The study was approved by the local Ethics Committee (165-2020; 93-2023bis) and was conducted in accordance with the Declaration of Helsinki. All patients signed written informed consent prior to participation.

At baseline (i.e., before implantation or battery replacement), patients were evaluated by a blinded neurologist using the Part III of the Movement Disorder Society-Unified Parkinson’s Disease Rating Scale (MDS-UPDRS-III), in Meds-OFF/Stim-OFF condition (after overnight withdrawal of all dopaminergic medication and at least 30 minutes after switching off STN-DBS, when present). After each two-week period (in cDBS or aDBS), the MDS-UPDRS-III evaluation was repeated in the best therapeutic condition (Meds-ON/Stim-ON, i.e., in the morning, 90 minutes after the usual dopaminergic medication and with stimulation switched on with the ongoing treatment). Clinical improvement in the two stimulation modes was defined as the percentual change with respect to the baseline score (%MDS-UPDRS-III).

Patients also completed the Hauser diary [62] for the last three consecutive days in each stimulation mode. From these data, we derived three self-reported motor measures: time spent in the OFF state (OFF), time spent in the ON state without troublesome dyskinesia (good-on-time [GOT], defined as ON time without or with mild dyskinesia), and time spent in ON state without any dyskinesia (ON_NoDysk_). Patients also recorded sleep/wake periods and medication intake times, which were used to determine patient-specific sleep and wake states. Self-reported motor measures were expressed as a percentage of wake time (%OFF, %GOT, and %ON_NoDysk_) and used as indicators of clinical benefit.

We assessed the significance of the clinical improvement (measured by %MDS-UPDRS-III) in both cDBS and aDBS mode using a one-sample t-test against zero. Then, differences in the %MDS-UPDRS-III between stimulation modes were assessed using a paired t-test. Moreover, we tested the self-perceived clinical difference between cDBS and aDBS (measured by %OFF, %GOT, and %ON_NoDysk_) using a paired t-test. Parametric t-tests were chosen after checking for data normality with the Shapiro-Wilk test. Multiple comparisons were corrected using the Holm-Bonferroni method. The effect size (es) was calculated using Cohen’s d. Statistical significance was set at p = 0.05.

Differences between stimulation modes for MDS-UPDRS-III and GOT were assessed also relative to the Minimal Clinically Important Difference (MCID) [63,64], which represents a change deemed clinically relevant by patients. Threshold for MCID were set at -3.25 points for MDS-UPDRS-III [65] and at 2 hours for GOT [66].

All analyses in the present manuscript were performed using MATLAB 2025a (The MathWorks Inc., Natick, MA, USA).

### Adaptive paradigm of the AlphaDBS device

cDBS was programmed according to standard clinical practice [67,68]. All patients received single-monopolar stimulation, with the active stimulation contact selected based on clinical monopolar review and kept unchanged between cDBS and aDBS (**Supplementary Table 1)**. In aDBS mode, stimulation amplitude was adjusted on a minute-by-minute basis according to the recorded STN-LFP amplitude following a linear proportional algorithm, while frequency and pulse width were held constant and matched to the cDBS mode.

For each patient, the recording contact pair was selected as the pair showing the most prominent beta peak during set-up recordings in the Meds-OFF/Stim-OFF condition and was subsequently verified for modulation after DBS activation.

Real-time LFP signal inspection during programming excluded the presence of cardiac artifacts [69]. Given the relatively short follow-up period of two weeks, it is likely that recordings remained free of major cardiac artifact contamination throughout the study.

Of note, the AlphaDBS system allows for the selection of any pair of recording contacts, provided they are not active for stimulation. The frequency range used to monitor STN-LFP amplitude was centered on each patient’s individually identified beta peak and was widened as needed based on individual recordings. The AlphaDBS device allows customization of the monitored frequency range, rather than limiting detection to a fixed 5 Hz bandwidth.

STN-LFP amplitude was recorded with one-second resolution, normalized to the total amplitude in the 5-34 Hz range, and smoothed with an exponential moving average with a time constant of 50 s. For aDBS programming, the clinician set two beta amplitude values (βmin and βmax) that defined the range within which the current varied linearly between a minimum and a maximum stimulation limit (Amin and Amax, **Figure 1**), chosen independently for the two hemispheres.

**Figure 1:**
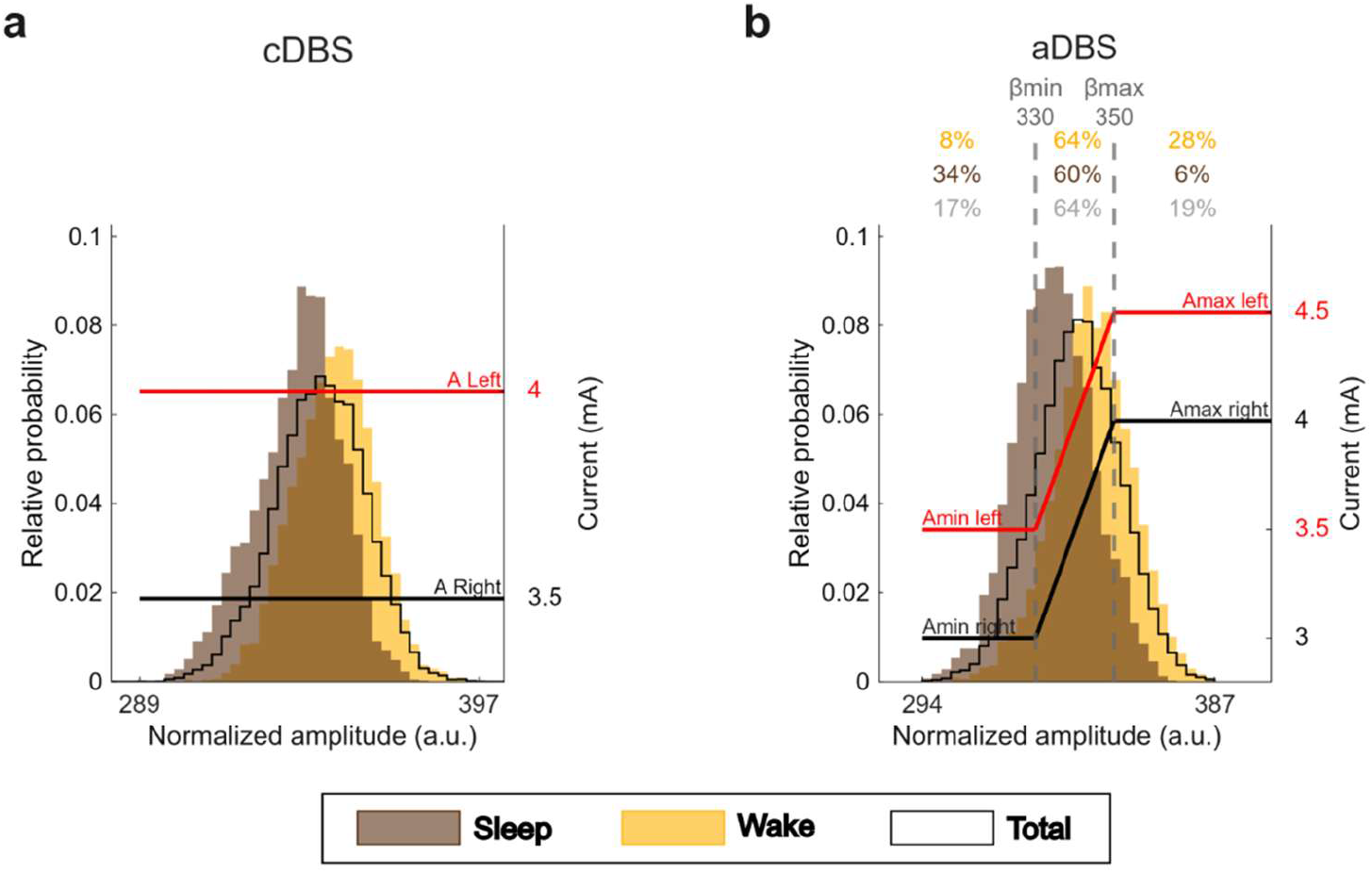
STN-LFP amplitude distribution and stimulation programming for one representative patient in cDBS and aDBS. (a) Probability distribution of the STN-LFP amplitude within a patient-specific range (12-19 Hz) collected with one-minute resolution over two weeks in cDBS for a representative patient (pt. 2). The histogram is separated into sleep (brown) and wake (yellow) states, while the distribution for the total time is shown by the black line. Red and black solid lines indicate the stimulation current set in cDBS for left and right STN, respectively. (b) Same as (a), but for aDBS. Vertical dotted lines indicate the biomarker thresholds for current adjustment (βmin and βmax). Red and black solid lines show the stimulation current at a specific reading for the left and right hemispheres, respectively. The current is modulated within a predefined, clinically effective range (Amin-Amax). Numbers at the top indicate the percentage of time the beta amplitude was below βmin, between βmin and βmax, and above βmax for the sleep (brown) and wake (yellow) states, as well as over the total recording time (grey). Abbreviations: a, adaptive; A, predefined, clinically effective amplitude; β, average normalized beta amplitude; c, conventional; DBS, deep brain stimulation; LFP, local field potentials; STN, subthalamic nucleus.

The two stimulation limits were defined as the minimum current amplitude (Amin) providing 40-50 % clinical benefit in Meds-OFF state (i.e., after overnight suspension of all dopaminergic drugs) and the maximum amplitude (Amax) in the absence of side effects in the Meds-ON condition (i.e., at 60 min after the intake of 100/25 mg levodopa/carbidopa) [48,49]. The device operates in a single-drive mode, in which one hemisphere serves as the driver controlling amplitude delivery to both STN.

To verify a sustained modulation of stimulation in aDBS, we calculated, for each day, the percentage of time during which the stimulation current varied by less than 0.2 mA for at least 60 consecutive minutes. A current variation inferior to 0.2 mA was arbitrarily defined as clinically irrelevant. Patients who exhibited such minimal current variation for more than 50% of their wake time were excluded from further analyses. To ensure comparability between the two stimulation modes, we calculated the total electrical energy delivered (TEED) in both conditions. In aDBS, TEED was computed for each one-minute current value. Median TEED values were first computed within each day and subsequently aggregated across days by taking the median of daily medians, separately for the sleep and wake state and for the total time. Impedances were considered constant and equal to 1 KΩ for all patients. Differences in TEED across conditions (cDBS and aDBS in sleep, wake, and total time) were assessed for the driver hemisphere using the Friedman test after checking for normality with the Shapiro-Wilk test. Post-hoc pairwise comparisons were performed using Wilcoxon signed-rank tests and corrected for multiple comparisons using the Holm-Bonferroni method. The effect size was calculated using Rank-Biserial Correlation [70]. Statistical significance was set at p = 0.05. To assess the clinical significance of differences in TEED, because frequency and pulse width remained unchanged between modes, we directly compared the fixed current amplitude in cDBS with the median current amplitude during the wake state in aDBS.

### Spectral analysis

The AlphaDBS device independently records two signals: i) the normalized beta amplitude within the patient-specific frequency range every minute, which serves as the input signal for current adaptation, ii) the amplitude spectrum in the 5-34 Hz range (1 Hz resolution, nV) every ten minutes. In the present manuscript, spectral analysis are performed exclusively on the second signal, which was recorded continuously for the two weeks in cDBS and aDBS.

From each raw amplitude spectrum we computed the power spectral density (PSD, nV^2^/Hz). Each PSD was then decomposed into its aperiodic and periodic components following the method described by Donoghue and coll. [71] (FOOOF algorithm with peak width limits = [2, 8], maximum number of peaks = 3, peak threshold = 1.5, aperiodic mode = ‘knee’; **Figure 2** and **Supplementary Figure 1**). The knee parameter was included in the model to account for the observed bending of the PSD in the log-log space at low frequencies. The goodness of each fit was evaluated through the squared coefficient of determination (R^2^) of the full model (sum of the aperiodic and periodic models in the logarithmic space), and fits with R^2^ values above 0.85 were kept for further processing [72].

**Figure 2:**
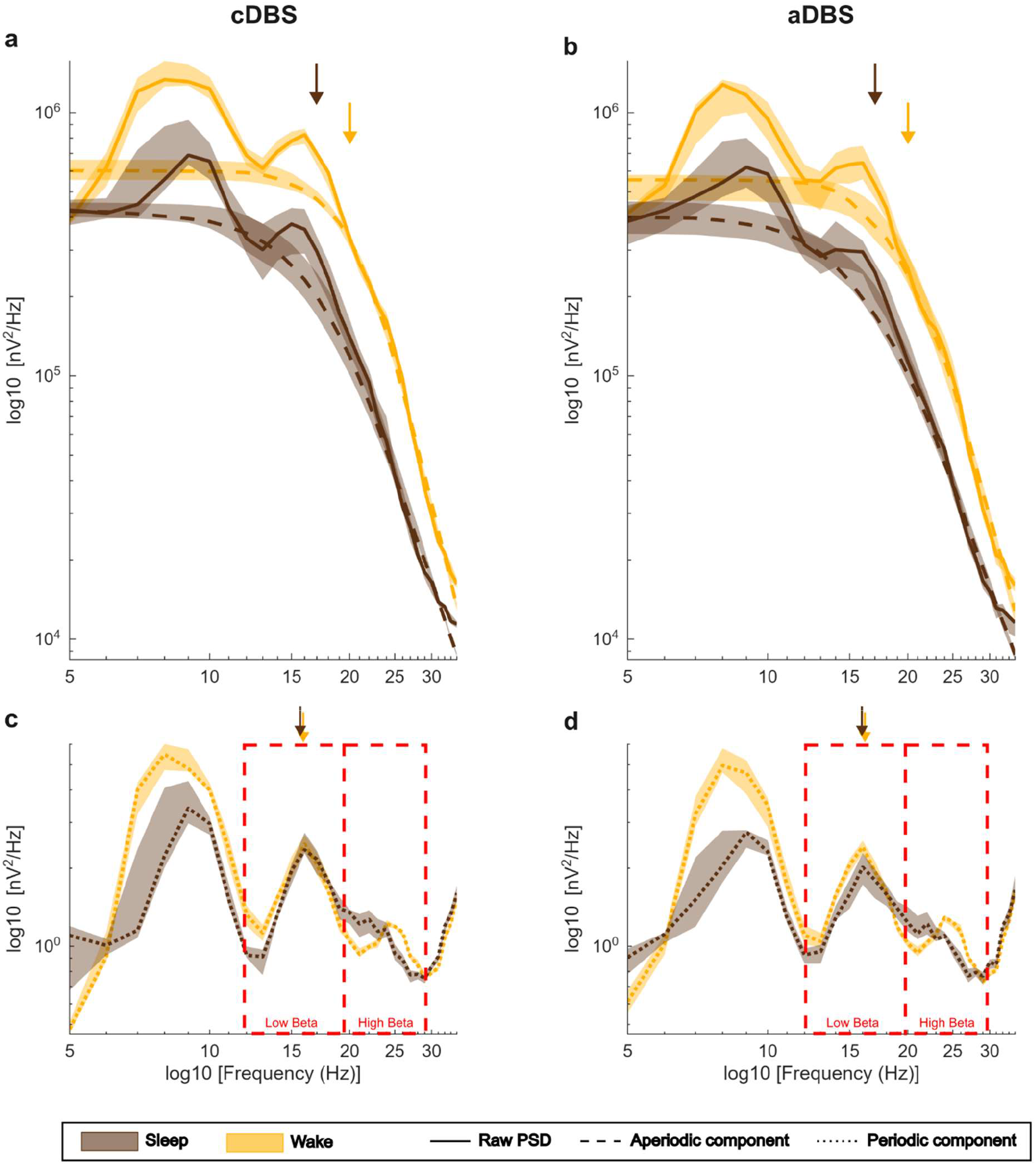
STN-LFP PSD with periodic and aperiodic component for one representative patient in cDBS and aDBS. (a) Loglog plot of median STN-LFP PSD (solid line) and median aperiodic component (dashed line) for one representative patient (pt. 2) in cDBS, separately for sleep (brown) and wake (yellow) states. Shaded regions correspond to the interquartile range (first to third quartile). Arrows represent the knee frequency for each state and mode (for pt. 2, sleep: 17 Hz and wake: 20 Hz). (b) Same as (a) but for aDBS (knee frequency for pt. 2, sleep: 17 Hz and wake: 20 Hz). (c) Loglog plot of median STN-LFP periodic component (dotted line) for one representative patient (pt. 2) in cDBS, separately for sleep (brown) and wake (yellow) states. Shaded regions correspond to the interquartile range (first to third quartile). Low beta and high beta bands are highlighted with red boxes. Arrows represent the patient-specific peak for each state and mode (for pt. 2, 16 Hz for sleep and wake). (d) Same as (c) but for aDBS patient-specific peak (for pt. 2, 16 Hz for sleep and wake). Abbreviations: a, adaptive; c, conventional; DBS, deep brain stimulation; LFP, local field potentials; PSD, power spectral density; STN, subthalamic nucleus.

For each state (sleep/wake) and mode (cDBS/aDBS), spectral peaks identified by the FOOOF algorithm were first clustered based on their frequency using the DBSCAN method (neighborhood search radius = 2, minimum number of neighboring points = 10 % of the total available points), followed by visual inspection. Then, for each cluster, the characteristic frequency of each cluster was selected as the mode of the frequencies of the peaks belonging to that cluster. Finally, for each state and mode, the most prominent characteristic peaks within the patient-specific frequency range used to modulate current delivery in aDBS (hereafter referred as patient-specific peak) was used as the center of a 5 Hz patient-specific range (hereafter referred as peak range) for subsequent analyses (**Figure 2** and **Supplementary Figure 1**). This approach ensured that the frequency ranges for power calculation were centered on meaningful spectral peaks and that the window width was standardized to 5 Hz across patients.

From each raw PSD, we calculated the power in the low beta band ([12–20] Hz, *rLBP*), the high beta band ([21–30] Hz, *rHBP*), and in the peak range (*rPP*), as the sum of the power values in the frequency range of interest.

The periodic components extracted with the FOOOF algorithm (i.e., the flattened PSD obtained subtracting in the log-log space the aperiodic component from the raw PSD) were first transformed from logarithmic to linear space. Then, we calculated the power in the low beta band (*pLBP*), the high beta band (*pHBP*), and in the peak range (*pPP*), as the sum of the power values in the frequency ranges of interest. The aperiodic activity was characterized using the aperiodic parameters extracted with the FOOOF algorithm (*Offset, Slope*, and *Knee*). Broadband power (*BP*), *Offset, Slope*, and knee frequency (*KneeFreq*) were used for further analysis, as these features have previously been identified in the literature as relevant for PD [53–55,73]. *BP* was calculated for each PSD as the sum of the aperiodic component in the logarithmic space from 5 to 34 Hz [55]. *KneeFreq* was calculated from the *Knee* and the *Slope* as *knee*^1/*slope*^ and represents approximately the frequency of the characteristic bend in the aperiodic 1/f^n^ PSD [74,75] (**Figure 2** and **Supplementary Figure 1**).

Overall, we identified ten spectral features of interest, divided in three classes: i) power of the raw PSD within each frequency bands of interest (*rLBP, rHBP*, and *rPP*); ii) power of the periodic components extracted with FOOOF in the same bands (*pLBP, pHBP*, and *pPP*); and iii) total power of the aperiodic component extracted with FOOOF (*BP*) and the features defining it (*Offset, Slope, KneeFreq*).

For each patient, state, mode and day, we computed the median and interquartile range of each spectral feature (noted as *x*_*med*_ and *x*_*iqr*_, given the spectral feature *x*). Finally, representative values for each patient were obtained as the median values across days (noted as *x*_*MED*_ and *x*_*IQR*_, given the spectral feature *x*).

For each spectral feature *x*, the effect of state and mode on *x*_*MED*_ and *x*_*IQR*_ was evaluated using a two-factor, within-subject repeated measures ANOVA. Normality of residuals was assessed using the Shapiro-Wilk test. When the normality assumption was violated, the data were transformed using the Aligned Rank Transform [76] prior to the ANOVA. Benjamini & Hochberg false discovery rate correction was applied separately for the three classes of features to each effect (state, mode and interaction). The effect size was calculated using the partial eta squared [77].

Subsequently, in case of significant interaction, single-effect analyses were performed using paired t-test. In case of data transformed using the Aligned Rank Transform for the repeated measures ANOVA, paired t-test was performed on aligned-and-ranked responses using ART-C procedure of ARTool to avoid inflating Type I error rates [78]. Holm-Bonferroni correction was applied separately within each feature and within each set of comparisons addressing the same effect (i.e., mode or state). The effect size was calculated using Cohen’s d. Statistical significance was set at p = 0.05.

### Linear mixed effects models

We built linear mixed-effects models [79] to describe the contribution of the periodic and aperiodic components and the stimulation mode to the difference in power of the raw PSD between sleep and wake states, in each spectral band: low beta band, high beta band, and peak range (Model 1, Equation 1).

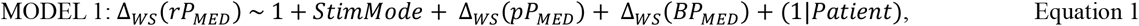

The dependent variable was the change in power of the raw PSD between sleep and wake (Δ_*ws*_ (*rP*_*MED*_)). Fixed effects included the stimulation mode (*StimMode*), the z-scored change in power of the periodic component between sleep and wake (Δ_*ws*_ (*pP*_*MED*_)), and the z-scored change in *BP* between sleep and wake (Δ_*ws*_ (*BP*_*MED*_)). Patients’ identity was included as a random intercept to account for between-subject variability.

To test the specific effects of the three aperiodic parameters, we developed a second model as follows (Model 2, Equation 2):

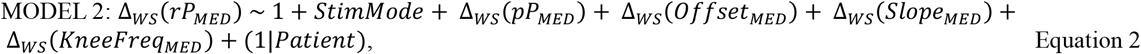

We replaced Δ_*ws*_ (*BP*_*MED*_) with the z-scored change in *Offset, Slope*, and *KneeFreq* between sleep and wake (Δ_*ws*_ (*Offset*_*MED*_), Δ_*ws*_ (*Slope*_*MED*_), *and* Δ_*ws*_ (*KneeFreq*_*MED*_); Equation 2).

To account for multiple comparisons across frequency bands, p-values for each fixed effect of interest were corrected across the three spectral bands (low beta, high beta, and peak range) using the Benjamini & Hochberg false discovery rate correction. Statistical significance was set at p = 0.05.

## RESULTS

### Participants selection

We enrolled eleven patients with idiopathic PD (**Table 1**). The first four patients had been stimulated for over three years with the Activa PC IPG (Medtronic Inc.) prior to enrollment, and received the AlphaDBS IPG at the time of battery replacement. The remaining seven patients received the AlphaDBS IPG as their first implant (de novo). Patients 1 to 8 were implanted with model 3389 leads (Medtronic Inc.), whereas patients 9 to 11 received model 6172 directional leads (Abbott) (**Table 1**). Electrode localization is shown in **Supplementary Figure 2**.

**Table 1:**
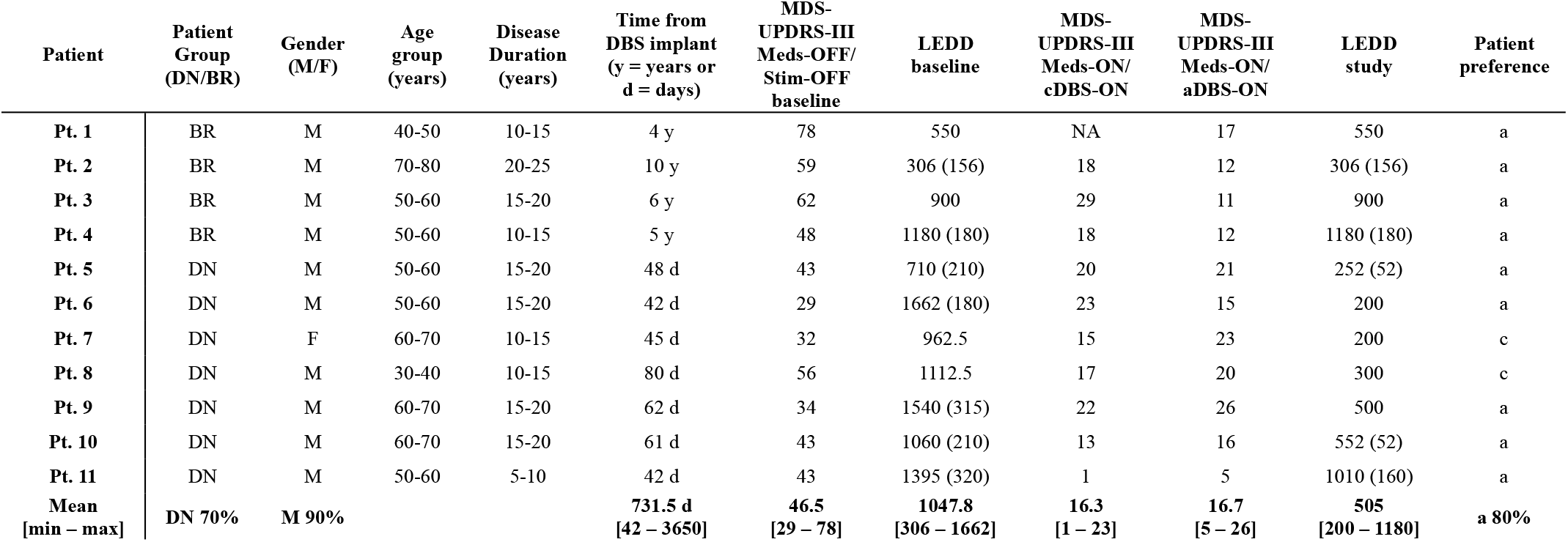
Demographic and clinical characteristics of the patients included in the study. Patients received the AlphaDBS device either at first implantation (de novo, DN) or at battery replacement (BR). Age, disease duration and time from surgery refer to the time of study start. MDS-UPDRS-III evaluations were performed at baseline either in the Meds-OFF state (i.e., after overnight withdrawal of all dopaminergic medication) if de novo, or in the Meds-OFF/Stim-OFF state (at least 30 minutes after switching off STN-DBS) if undergoing battery replacement, and after two weeks in best medical condition (Meds-ON/Stim-ON, i.e., in the morning, 90 minutes after the usual morning medication and with stimulation switched on). LEDD was calculated based on the pharmacological therapy at baseline and during the two-week period in each stimulation mode (study). Values in parentheses indicate LEDD for dopamine agonists only. Medications remained unchanged between cDBS and aDBS. The last column reports the stimulation mode blindly preferred by each patient. Patient 3 was excluded from the analysis, therefore the reported mean [min – max] do not include this patient. Abbreviations: a, adaptive; BR, battery replacement; c, conventional; d, days; DBS, deep brain stimulation; DN, de novo; F, female; LEDD, levodopa equivalent daily dose; M, male; pt., patient; y, years.

| Patient | Patient Group (DN/BR) | Gender (M/F) | Age group (years) | Disease Duration (years) | Time from DBS implant (y = years or d = days) | MDS-UPDRS-III Meds-OFF/ Stim-OFF baseline | LEDD baseline | MDS-UPDRS-III Meds-ON/ cDBS-ON | MDS-UPDRS-III Meds-ON/ aDBS-ON | LEDD study | Patient preference |
| --- | --- | --- | --- | --- | --- | --- | --- | --- | --- | --- | --- |
| Pt. 1 | BR | M | 40-50 | 10-15 | 4 y | 78 | 550 | NA | 17 | 550 | a |
| Pt. 2 | BR | M | 70-80 | 20-25 | 10 y | 59 | 306 (156) | 18 | 12 | 306 (156) | a |
| Pt. 3 | BR | M | 50-60 | 15-20 | 6 y | 62 | 900 | 29 | 11 | 900 | a |
| Pt. 4 | BR | M | 50-60 | 10-15 | 5 y | 48 | 1180 (180) | 18 | 12 | 1180 (180) | a |
| Pt. 5 | DN | M | 50-60 | 15-20 | 48 d | 43 | 710 (210) | 20 | 21 | 252 (52) | a |
| Pt. 6 | DN | M | 50-60 | 15-20 | 42 d | 29 | 1662 (180) | 23 | 15 | 200 | a |
| Pt. 7 | DN | F | 60-70 | 10-15 | 45 d | 32 | 962.5 | 15 | 23 | 200 | c |
| Pt. 8 | DN | M | 30-40 | 10-15 | 80 d | 56 | 1112.5 | 17 | 20 | 300 | c |
| Pt. 9 | DN | M | 60-70 | 15-20 | 62 d | 34 | 1540 (315) | 22 | 26 | 500 | a |
| Pt. 10 | DN | M | 60-70 | 15-20 | 61 d | 43 | 1060 (210) | 13 | 16 | 552 (52) | a |
| Pt. 11 | DN | M | 50-60 | 5-10 | 42 d | 43 | 1395 (320) | 1 | 5 | 1010 (160) | a |
| Mean [min – max] | DN 70% | M 90% |  |  | 731.5 d [42 – 3650] | 46.5 [29 – 78] | 1047.8 [306 – 1662] | 16.3 [1 – 23] | 16.7 [5 – 26] | 505 [200 – 1180] | a 80% |

Patient 3 was excluded from the analysis because current modulation in aDBS during the wake state was not sustained (the stimulation current varied by less than 0.2 mA for at least 60 consecutive minutes for 76.8 % of the wake time, see Methods, **Supplementary Figure 3** and **Supplementary Table 2**).

### cDBS and aDBS show comparable clinical efficacy

cDBS and aDBS resulted in significant clinical improvements compared with baseline assessments, with %MDS-UPDRS-III scores improving by 59.1 [20.7 – 97.7] % in cDBS and by 59.9 [23.5 – 88.4] % in aDBS (t-test p-values and Cohen’s d es. cDBS: p < 0.001, es = 2.66, and aDBS: p < 0.001, es = 2.73; **Table 1**). All numeric results are reported as *average across patients [range]*, unless differently specified. Effect sizes are reported only for significant comparisons. The improvement was not significantly different between cDBS and aDBS (t-test, p = 0.83).

Regarding the clinical evaluations derived from the Hauser diary (see Methods), %OFF was lower in aDBS compared with cDBS, (cDBS: 31.5 [0 – 80] % and aDBS: 13.6 [0 – 70] %; **Supplementary Table 3**), while %GOT and %ON_NoDysk_ were larger in aDBS compared with cDBS (%GOT – cDBS: 66.7 [20 – 96.8] % and aDBS 86.3 [30 – 100] %, %ON_NoDysk_ – cDBS: 52.8 [1 – 89.1] % and aDBS: 71.8 [10.9 – 100] %; **Supplementary Table 3**). However, the difference between the two stimulation modes was never statistically significant due to the large inter-patient variability (t-test, %OFF: p = 0.18, %GOT: p = 0.25, and %ON_NoDysk_: p = 0.25; **Supplementary Table 3**).

The difference in MDS-UPDRS-III between cDBS and aDBS reached the MCID (see Methods) in one patient in favor of cDBS (pt. 7; **Table 1**) and in three patients in favor of aDBS (pt. 2, 4, 6; **Table 1**). For GOT, the difference between cDBS and aDBS reached the MCID in two patients in favor of cDBS (pt. 2 and 7; **Supplementary Table 3**) and in five patients in favor of aDBS (pt. 1, 5-6, 9-10; **Supplementary Table 3**). Notably, under blinded conditions, eight out of ten patients expressed a preference for aDBS at the end of the study (pt. 1-6, 9-11; **Table 1**).

### Stimulation was reduced during sleep in aDBS

Anecdotal reports from several patients describing improved sleep quality with DBS motivated the analyses presented in this study. Timing for sleep and wake states are displayed in **Supplementary Table 4**.

Detailed programming parameters and TEED values for cDBS and aDBS (for the latter, analyzed also during sleep and wake states separately) are provided in **Supplementary Table 1** (cDBS: 56.6 [15.3 – 140] µW, aDBS – sleep: 48.2 [5 – 117.6] µW, wake: 59.7 [9.4 – 147.9] µW, and total time: 56.7 [6.3 – 147.9] µW). TEED in aDBS was significantly higher in the wake compared with the sleep state (Wilcoxon signed-rank test p = 0.023, Rank-Biserial Correlation es = 0.93; **Supplementary Table 1**). TEED in aDBS was comparable to that in cDBS during both the wake state and the total time (Wilcoxon signed-rank test, p = 0.47 and 0.95, respectively). In contrast, TEED in aDBS during sleep was significantly lower than that in cDBS (Wilcoxon signed-rank test p = 0.041, Rank-Biserial Correlation es = 0.86; **Supplementary Table 1**).

### Patient-specific frequency peak remains stable across states and modes

For all patients in all states and modes, STN-LFP PSD showed two or three peaks distributed in the 5-34 Hz range (see Methods). Patient-specific frequency range used to modulate current delivery in aDBS were predominantly within the low beta band. In four patients, the lower bound extended into the alpha band, while in one patient the upper bound reached the high beta band (**Supplementary Table 1**). Consequently, patient-specific peaks were all located in the low beta band, except for one peak in aDBS during the sleep state (pt. 9: 11 Hz; **Table 2**). The frequency of patient-specific peaks remained stable across states and modes for all patients (peak frequency difference wake-sleep: 0.4 [0 – 2] Hz in cDBS and 0.7 [0 – 3] Hz in aDBS; peak frequency difference aDBS-cDBS: -0.3 [-1 – 0] Hz in sleep and 0 [0 – 0] Hz in wake; **Table 2** and **Supplementary Figure 1**).

**Table 2:** Frequency peak characterization. Frequency (Hz) of the patient-specific peaks in each state (sleep/wake) and mode (cDBS/aDBS). Abbreviations: a, adaptive; c, conventional, DBS, deep brain stimulation; pt., patient.

| Patient | cDBS<br>Sleep | cDBS<br>Wake | aDBS<br>Sleep | aDBS<br>Wake |
| --- | --- | --- | --- | --- |
| Pt. 1 | 13 | 13 | 12 | 13 |
| Pt. 2 | 16 | 16 | 16 | 16 |
| Pt. 4 | 12 | 13 | 12 | 13 |
| Pt. 5 | 13 | 14 | 13 | 14 |
| Pt. 6 | 12 | 12 | 12 | 12 |
| Pt. 7 | 16 | 16 | 16 | 16 |
| Pt. 8 | 20 | 20 | 20 | 20 |
| Pt. 9 | 12 | 14 | 11 | 14 |
| Pt. 10 | 17 | 17 | 16 | 17 |
| Pt. 11 | 16 | 16 | 16 | 16 |

### Sleep decrease in beta power is a combination of aperiodic component and oscillatory activity

Representative median power values (*x*_*MED*_) of raw PSD in all spectral bands (*rLBP*_*MED*_ *rHBP*_*MED*_, and *rPP*_*MED*_, see Methods) were significantly larger in the wake compared with the sleep state (repeated measures ANOVA p-values and partial eta squared es. *rLBP*_*MED*_ : p = 0.0024, es = 0.69, **Figure 3a**, *rHBP*_*MED*_: p = 0.0015, es = 0.76, **Figure 3b**, and *rPP*_*MED*_: p = 0.0033, es = 0.64, **Figure 3c; Supplementary Table 5**).

**Figure 3:**
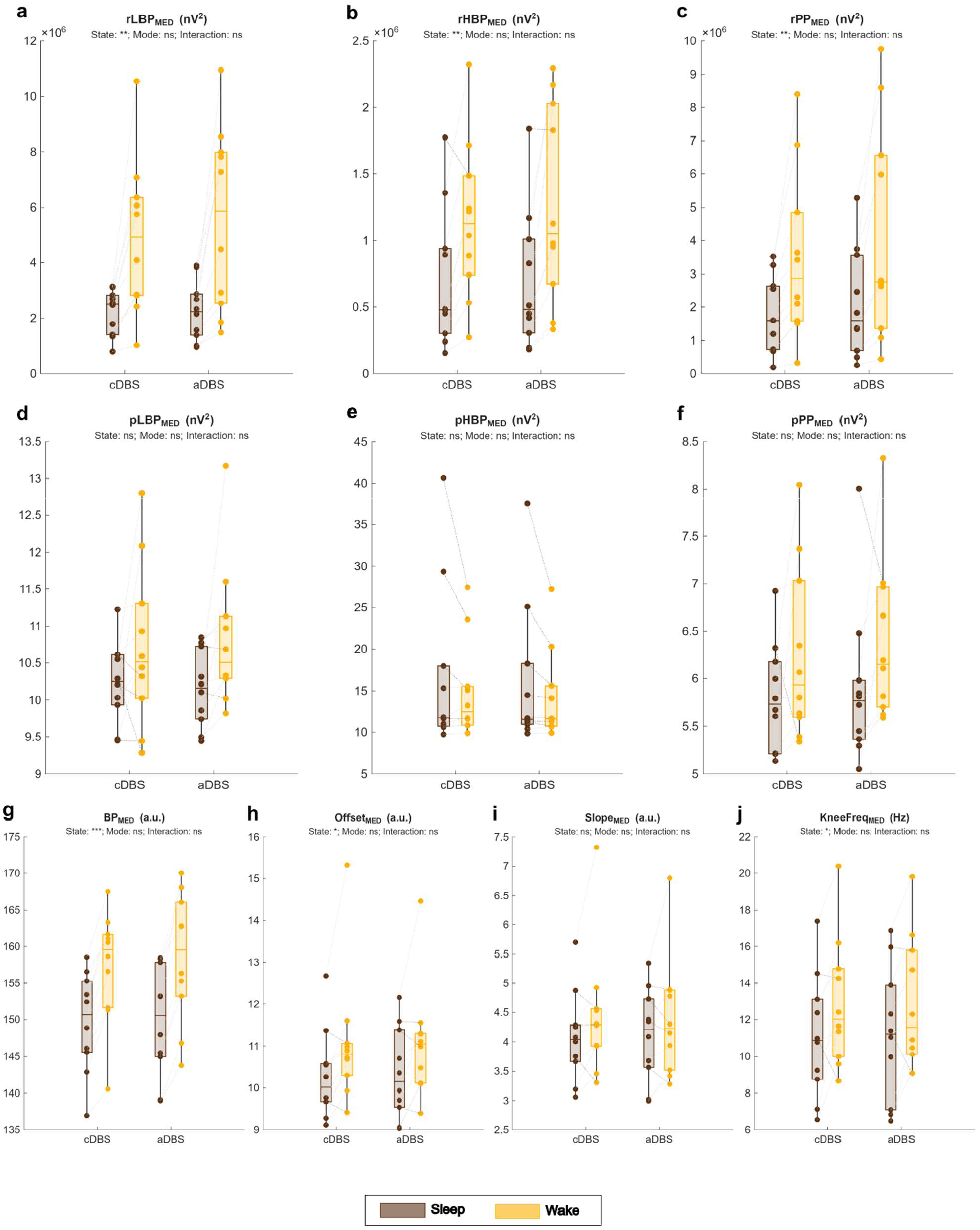
Effect of state and mode on each spectral feature (median). For each spectral feature x, box chart of x_MED_ in cDBS and aDBS separately for the sleep (brown) and wake (yellow) state. Each dot represents the x_MED_ of an individual patient in the corresponding state and mode and grey dotted lines connect the same patient between sleep and wake state (light grey if wake > sleep and dark grey otherwise). The subtitle repots the results of the repeated measures ANOVA test with Benjamini & Hochberg false discovery rate correction (p>0.05: ns, 0.01<p<0.05: *, 0.001<p<0.01: **, and p<0.001: ***). Abbreviations: a, adaptive; BP_MED_, overall median broadband power of the aperiodic component; c, conventional; DBS, deep brain stimulation; KneeFreq_MED_, overall median knee frequency of the aperiodic component; Offset_MED_, overall median offset of the aperiodic component; pHBP_MED_, overall median power of periodic component in the high beta band; pLBP_MED_, overall median power of periodic component in the low beta band; pPP_MED_, overall median power of periodic component in the peak range; rHBP_MED_, overall median power of raw PSD in the high beta band; rLBP_MED_, overall median power of raw PSD in the low beta band; rPP_MED_, overall median power of raw PSD in the peak range; Slope_MED_, overall median slope of the aperiodic component.

However, the power of periodic components in the same spectral bands (*pLBP*_*MED*_, *pHBP*_*MED*_, and *pPP*_*MED*_) did not differ significantly between sleep and wake state (repeated measures ANOVA p-values. *pLBP*: p = 0.14, **Figure 3d**, *pHBP*: p = 1, **Figure 3e**, and *pPP*_*MED*_ : p = 0.14, **Figure 3f**; **Supplementary Table 5**). Notably, for *pPP*_*MED*_, even though the comparison was not statistically significant, all patients except one showed higher periodic power during wake compared to sleep in both cDBS and aDBS (**Figure 3f** and **Supplementary Table 5**).

Instead, the total power of the aperiodic component *BP*_*MED*_, and two of the features determining it (*Offset*_*MED*_, and *KneeFreq*_*MED*_) were significantly larger in the wake compared with the sleep state (repeated measures ANOVA p-values and partial eta squared es. *BP*_*MED*_: p < 0.001, es = 0.90, **Figure 3g**, *Offset*_*MED*_: p = 0.031, es = 0.49, **Figure 3h**, and *KneeFreq*_*MED*_: p = 0.031, es = 0.45, **Figure 3j; Supplementary Table 5**).

Mode factor was marginally significant only for *pHBP*_*MED*_, with cDBS being larger than aDBS (repeated measures ANOVA p = 0.054, partial eta squared es = 0.48, **Figure 3e** and **Supplementary Table 5**).

*x*_*MED*_ did not show significant interaction between state and mode for any spectral feature (**Figure 3** and **Supplementary Table 5**).

### Beta power of raw PSD is more variable in the wake state independently of the stimulation mode

Representative interquartile range of power values (*x*_*IQR*_) of raw PSD in all spectral bands (*rLBP*_*IQR*_, *rHBP*_*IQR*_, and *rPP*_*IQR*_, see Methods) were significantly larger in the wake compared with the sleep state (repeated measures ANOVA p-values and partial eta squared es. *rLBP*_*IQR*_: p = 0.002, es = 0.75, **Supplementary Figure 4a**, *rHBP*_*IQR*_: p = 0.014, es = 0.50, **Supplementary Figure 4b**, and *rPP*_*IQR*_ : p = 0.014, es = 0.52, **Supplementary Figure 4c; Supplementary Table 6**). *rHBP*_*IQR*_ showed significant interaction between state and mode (repeated measures ANOVA p = 0.019 and partial eta squared es = 0.58). Post hoc analysis showed that *rLBP*_*IQR*_ was significantly larger in the wake compared to the sleep state in both cDBS and aDBS (paired t-test p-values and Cohen’s d es. cDBS: p < 0.001, es = 1.88, and aDBS: p < 0.001, es = 1.81, **Supplementary Figure 4a** and **Supplementary Table 6**). However, *rLBP*_*IQR*_ did not significantly differ between cDBS and aDBS in neither sleep or wake state (paired t-test p = 0.88 in both sleep and wake states, **Supplementary Figure 4a** and **Supplementary Table 6**). Averaging across patients, the difference in *rLBP*_*IQR*_ between wake and sleep was larger in aDBS compared to cDBS (Δ (*rLBP*_*IQR*_) = 1263583.5 nV^2^ in cDBS, and 1746760.1 nV^2^ in aDBS, +38.2%).

All other spectral features did not show significant interaction between state and mode, nor significant effect for mode or state factors (**Supplementary Figure 4** and **Supplementary Table 6**).

### Aperiodic component largely accounts for sleep-wake differences in the raw PSD

Linear mixed-effects models clarified the contribution of periodic and aperiodic component on the difference in the raw PSD between sleep and wake.

Model 1, taking into account the total aperiodic component (see Methods), showed significant effect of the intercept and Δ_*ws*_(*BP*_*MED*_) for all frequency bands (**Table 3**), and of Δ _*ws*_ (*pP*_*MED*_) for low beta band and peak range (**Table 3**). Estimates of Δ _*ws*_ (*BP*_*MED*_) were always larger compared with estimates of Δ _*ws*_ (*pP*_*MED*_) (+30.1% for low beta band and +69.5% for peak range; **Table 3**). The effect of stimulation mode (*StimMode*) was not significant. The coefficients of determination of the models were 0.98, 0.97, and 0.93 for low beta band, high beta band, and peak range, respectively (**Figure 4a-c and Table 3**).

**Table 3.** Linear mixed-effects models. For each model (Model 1 and 2, in upper and lower table, respectively) and frequency band (low beta band, high beta band, and peak range), estimates and p-values for each effect. Abbreviations: BP_MED_, overall median broadband power of the aperiodic component; Δ, delta wake-sleep; KneeFreq_MED_, overall median knee frequency of the aperiodic component; Offset_MED_, overall median offset of the aperiodic component; pP_MED_, overall median power of periodic component in the low beta band (pLBP_MED_), high beta band (pHBP_MED_), and in the peak range (pPP_MED_); rP_MED_, overall median power of raw PSD in the low beta band (rLBP_MED_), high beta band (rHBP_MED_), and in the peak range (rPP_MED_); R^2^ _adjusted_, coefficient of determination of the model; Slope, _MED_ overall median slope of the aperiodic component; StimMode, stimulation mode.

| Model 1 | $\Delta_{WS}(rLBP_{MED})$<br>($R^2_{adjusted} = 0.98$) | | $\Delta_{WS}(rHBP_{MED})$<br>($R^2_{adjusted} = 0.97$) | | $\Delta_{WS}(rPP_{MED})$<br>($R^2_{adjusted} = 0.93$) | |
| --- | --- | --- | --- | --- | --- | --- |
|  | Estimate | p-value | Estimate | p-value | Estimate | p-value |
| (Intercept) | 3537688.34 | < 0.001 | 484712.65 | < 0.001 | 2206680.006 | < 0.001 |
| StimMode | -358142.045 | 0.33 | 17793.35 | 0.71 | -205409.12 | 0.62 |
| $\Delta_{WS}(pP_{MED})$ | 1413345.16 | < 0.001 | 82098.66 | 0.30 | 857742.46 | 0.015 |
| $\Delta_{WS}(BP_{MED})$ | 1839615.021 | < 0.001 | 333384.87 | < 0.001 | 1453457.63 | < 0.001 |

**Table 3. Linear mixed-effects models.**
| Model 2 | $\Delta_{WS}(rLBP_{MED})$<br>($R^2_{adjusted} = 0.94$) | | $\Delta_{WS}(rHBP_{MED})$<br>($R^2_{adjusted} = 0.95$) | | $\Delta_{WS}(rPP_{MED})$<br>($R^2_{adjusted} = 0.82$) | |
| --- | --- | --- | --- | --- | --- | --- |
|  | Estimate | p-value | Estimate | p-value | Estimate | p-value |
| (Intercept) | 3297957.75 | < 0.001 | 436965.40 | 0.0021 | 2058415.80 | 0.0023 |
| StimMode | -198321.61 | 0.75 | 49624.86 | 0.75 | -106566.31 | 0.75 |
| $\Delta_{WS}(pP_{MED})$ | 1423260.50 | 0.0038 | 25192.58 | 0.75 | 782910.19 | 0.019 |
| $\Delta_{WS}(Offset_{MED})$ | 7679070.24 | < 0.001 | 1434915.58 | < 0.001 | 6200981.48 | < 0.001 |
| $\Delta_{WS}(Slope_{MED})$ | -7306298.40 | < 0.001 | -1407719.35 | < 0.001 | -6152789.36 | < 0.001 |
| $\Delta_{WS}(KneeFreq_{MED})$ | 25562.071 | 0.96 | 151623.46 | 0.39 | 251147.68 | 0.78 |

**Figure 4:**
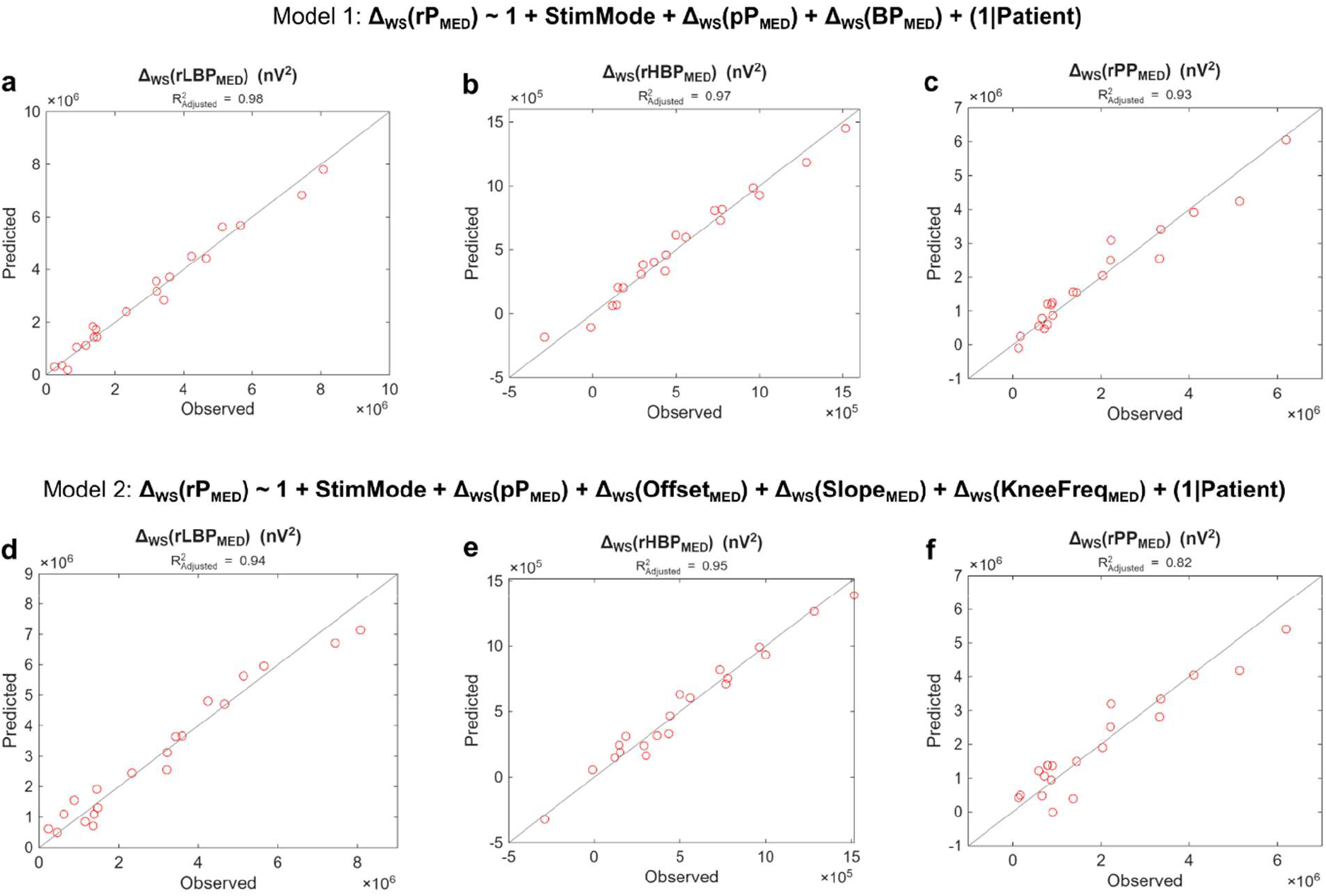
Linear mixed-effects models. (a) Plot of observed versus predicted values for Model 1 and low beta band. Each plot has 20 dots, i.e., 10 patients in 2 stimulation modes (cDBS/aDBS). Solid black line is the identity line. The subtitle indicates the coefficient of determination of the model (R^2^_adjusted_). (b) Same as (a) for high beta band. (c) Same as (a) for peak range. (d) Same as (a) for Model 2. (e) Same as (b) for Model 2. (f) Same as (c) for Model 2. Abbreviations: a, adaptive; BP_MED_, overall median broadband power of the aperiodic component; c, conventional; DBS, deep brain stimulation; Δ, delta wake-sleep; KneeFreq_MED_, overall median knee frequency of the aperiodic component; Offset_MED_, overall median offset of the aperiodic component; pP_MED_, overall median power of periodic component in the low beta band (pLBP_MED_), high beta band (pHBP_MED_), and in the peak range (pPP_MED_); rP_MED_, overall median power of raw PSD in the low beta band (rLBP_MED_), high beta band (rHBP_MED_), and in the peak range (rPP_MED_); Slope_MED_, overall median slope of the aperiodic component; StimMode, stimulation mode.

Model 2, decomposing the aperiodic component in its factors (see Methods), showed significant effect of intercept, Δ_*ws*_ (*Offset*_*MED*_), and Δ _*ws*_ (*Slope*_*MED*_) for all frequency bands, and of Δ _*ws*_ (*pP*_*MED*_) for low beta band and peak range (**Table 3**). Estimate of Δ _*ws*_ (*Offset*_*MED*_), and Δ _*ws*_ (*Slope*_*MED*_) were comparable and always larger compared with estimates of Δ _*ws*_ (*pP*_*MED*_) (Δ _*ws*_ (*Offset*_*MED*_): +439.5% for low beta band and +692% for peak range, Δ _*ws*_ (*Slope*_*MED*_): +413.4% for low beta band and +685.9% for peak range; **Table 3**). The effect of stimulation mode (*StimMode*) was not significant. The coefficients of determination of the models were 0.94, 0.95, and 0.82 for low beta band, high beta band, and peak range, respectively (**Figure 4d-f and Table 3**).

## DISCUSSION

The availability of new IPGs capable of chronically recording subcortical brain signals during daily life activities is expanding our understanding of PD pathophysiology and the mechanism of action of DBS. This major technological advancement has opened new perspectives for personalized neuromodulation therapies.

In this study, we investigated how STN oscillatory and aperiodic activity are modulated across the sleep-wake cycle in patients with PD undergoing either conventional or adaptive DBS.

First, we showed the stability of the patient-specific beta peak across states (sleep/wake) and modes (cDBS/aDBS) (see Methods and **Supplementary Figure 1**). This finding is particularly relevant for the effective programming of aDBS using currently available devices, which require the identification of a patient-specific frequency range for chronic monitoring. A substantial shift in the peak frequency between sleep and wake could result in prolonged stimulation at either the minimum or maximum amplitude level (Amin or Amax). It should also be noted that the peak stability described in our study primarily refers to the beta peak used to configure the aDBS mode, whose prominence may be attenuated and less clearly detectable during chronic active DBS [47,80]. In addition, several patients exhibited more than one oscillatory component within the PSD. Future studies should investigate in greater detail the temporal evolution and modulation of oscillatory activity, ideally through repeated recordings obtained after DBS washout periods of different durations. However, such approaches remain difficult to implement in clinical practice because of the rapid and often dramatic re-emergence of motor symptoms following stimulation withdrawal.

As a second main result, we observed a global reduction in raw STN spectral power during sleep across all investigated frequency bands (i.e., low beta band, high beta band, and peak range). This observation is consistent with previous literature [81] and extends these observations to a different sensing-enabled IPG platform. Previous sleep studies in patients with PD implanted for STN-DBS have consistently reported reduced beta power during NREM sleep and comparable or even higher beta power during REM sleep relative to wakefulness [14,81–83], thereby paving the way for neural decoding of sleep stages [84,85]. Moreover, beta power during NREM sleep has been positively correlated with measures of sleep fragmentation [86,87], highlighting its potential as a biomarker for sleep-aware aDBS algorithms. Despite these promising findings, previous studies relied exclusively on raw spectral power, thereby conflating the contributions of periodic and aperiodic activity. By leveraging the capability of the AlphaDBS device to chronically record the STN spectrum, rather than only the power within a patient-specific frequency range, we were able to disentangle these two components and investigate their respective contributions to sleep-related beta power suppression. Interestingly, we found that, at the group level, the power of the periodic component was not significantly reduced during sleep in any of the investigated frequency bands. In contrast, aperiodic broadband power showed a marked and consistent reduction during sleep, driven by changes in both the offset and the knee of the aperiodic component. We also observed that this sleep-wake modulation was not influenced by stimulation mode, whether cDBS or aDBS.

The linear mixed-effects models provided further mechanistic insight into the observed sleep-wake modulation of STN activity. Although repeated-measures ANOVA revealed no significant main effect of state on periodic beta power, the mixed models showed that changes in periodic beta activity within the low beta band and peak range still contributed to explaining variations in raw spectral power when jointly considered with aperiodic components. However, the contribution of aperiodic features, particularly broadband power, offset, and slope, was consistently larger than that of periodic activity, indicating that sleep-wake modulation of STN beta power is predominantly driven by broadband spectral shifts. Importantly, substantial inter-individual variability was observed. While some patients exhibited a marked suppression of beta oscillatory peaks during sleep (e.g., pt. 5 and 9; **Supplementary Figure 1**), others showed only modest changes or even the opposite pattern. In contrast, suppression of aperiodic features was robust and consistently observed across patients. This greater consistency likely explains why periodic activity contributed to the models to a lesser extent than aperiodic components. Larger cohorts will be needed to clarify whether true oscillatory suppression of STN beta activity occurs during sleep in a subset of patients with PD.

These findings closely align with emerging evidence in sleep neurophysiology, where cortical background activity is viewed as an indicator of arousal. Cortical studies in healthy subjects have shown that sleep is associated with reduced alpha and beta activity and increased slow-wave power [88] compared with wakefulness. Moreover, the aperiodic slope has been proposed as a marker of arousal, with steeper slopes observed in deeper sleep states [56–60]. This is consistent with theoretical models linking the aperiodic slope to the synaptic excitation/inhibition (E/I) balance [89], reflecting increased inhibition during sleep. A rat model of PD [90] further supported the association between the aperiodic slope and the E/I balance within the STN, particularly in the gamma band (30-100 Hz). In our dataset, however, we did not observe a significant sleep-wake modulation of the aperiodic slope. This absence may be explained by the lower frequency range examined in our study (12-30 Hz), compared with the higher frequencies typically investigated in previous studies (>30 Hz [89,90]).

Aperiodic offset, on the other hand, has been associated with neuronal population spiking [91] and global firing rates, which is consistent with the reduction in STN offset and broadband power observed during sleep. These findings suggest a state-dependent modulation of STN activity, likely reflecting global shifts in population activity rather than purely oscillatory (de-)synchronization. This interpretation is further supported by previous studies reporting decreased STN spiking frequency during sleep, associated with more irregular and bursting firing patterns [92].

Importantly, aperiodic parameters differ between cortical and deep brain structures [93], limiting the direct translation of cortical findings to the STN and warranting further investigation.

The similarity in findings between the low beta band and the peak range is expected, since mostly all patient-specific peaks fell within the low beta band. However, the absence of a significant periodic contribution to the linear mixed-effects model for the high beta band supports the notion of a functional dissociation between low and high beta bands in PD. Previous works have suggested that low beta is more closely linked to pathological synchrony, motor impairment, and clinical improvement with levodopa administration [94–99]. In contrast, high beta has been associated with distinct functional roles, including sensorimotor processing and movement-related dynamics [99–101], and it has been associated to specific modulation by DBS [95]. The lack of a significant periodic effect in the high beta in the present study is therefore consistent with the idea that sleep-wake spectral differences predominantly affect beta activity more tightly coupled to pathological network dynamics in PD, possibly reflecting the reduction of akinetic-rigid symptoms during sleep.

A crucial consideration in aDBS programming is how to effectively leverage the reduction of beta power during sleep. This phenomenon not only reflects the reduced motor demand of the resting state, but may also help prevent nocturnal stimulation-related side effects, including sleep deterioration [50]. Furthermore, reducing nocturnal energy delivery may mitigate the development of long-term habituation to therapy for certain symptoms, such as tremor [51]. The patient-specific histogram distribution across sleep and wake states should be considered when defining βmin (**Figure 1**). Setting βmin at higher levels enables continuous low-amplitude stimulation (Amin) during sleep and greater current modulation during wakefulness. Conversely, lower βmin values allow continuous modulation across the 24-hour cycle, although polarized toward two predominant stimulation amplitudes corresponding to the median values of the sleep and wake distributions (**Figure 1**). Consistently, we found that stimulation amplitude in aDBS was significantly reduced during sleep, while cDBS amplitude was comparable to that delivered by aDBS during wakefulness, indicating that the adaptive algorithm effectively tracks circadian state transitions in addition to pathological oscillatory activity. In the future, combining chronic STN-LFP recordings with polysomnography (PSG) may enable the identification of sleep stage-specific neural signatures and support the development of more refined adaptive algorithms [82,83,85].

Exploratory analyses did not reveal significant correlations between spectral features (raw, periodic, or aperiodic) and clinical measures (MDS-UPDRS-III or Hauser diary scores) during wakefulness in either cDBS or aDBS. Although this contrasts with the well-established association between beta power and motor symptom severity in PD [102–105], it is consistent with recent evidence suggesting that such correlations are highly variable and may be overestimated in small cohorts [55]. The same study [55] showed that aperiodic STN offset and broadband power were associated with motor symptom severity and dopamine depletion, respectively, supporting the inclusion of aperiodic spectral features as additional biomarkers for aDBS. However, most previous studies [55,102,104] were performed in acute post-operative settings without active DBS. We therefore hypothesize that chronic aDBS may suppress the pathological component of subthalamic beta activity while preserving physiological STN dynamics, potentially explaining the absence of clinical correlations in our cohort. This effect may have been further compounded by the mismatch between point-in-time clinical assessments and chronic at-home neural recordings.

From a neuromodulation perspective, our findings have direct implications. Current beta-driven aDBS algorithms operating on raw power measures may respond to broadband changes associated with arousal rather than exclusively to pathological beta oscillations. Oscillatory bursts represent temporally structured, low-entropy states that may constrain information processing, whereas aperiodic activity reflects higher-entropy, more desynchronized states available for information processing [106]. Integrating aperiodic metrics into adaptive control schemes could therefore help disentangle physiological sleep-related fluctuations in broadband activity from pathological beta (de-)synchronization and improve aDBS specificity and efficacy. Recent sleep-aware adaptive paradigms, including cortical discriminators [107,108] and stimulation adjustments across specific sleep stages [109], support the feasibility of circadian-informed neuromodulation. Our results suggest that subthalamic aperiodic features could complement these approaches and fit with the broader framework of digital chronotherapy in DBS [110]. Lastly, monitoring aperiodic activity may be particularly relevant in patients without a clear beta peak, potentially enabling sleep-related stimulation reduction even in the absence of a distinct oscillatory biomarker.

Our study has some limitations. First, the lack of sleep recording with PSG prevented the differentiation between REM and NREM sleep stages, which are known to exhibit distinct STN dynamics [82,83]. In addition, clinical sleep scales were not systematically collected, as sleep outcomes were not part of the original study objectives [5], although several patients spontaneously reported subjective sleep improvement during DBS.

Second, the wake state included heterogeneous behavioral and medication conditions, making it difficult to completely exclude the influence of motor and non-motor activities occurring during daily life [111–113]. Future home-monitoring approaches integrating behavioral recordings may help clarify STN modulation under real-world conditions.

Third, recordings were obtained under active DBS, meaning that pathological oscillations were likely already partially suppressed. Recordings acquired before IPG activation or during DBS washout periods may help better characterize intrinsic sleep-wake STN dynamics. Moreover, we could not investigate frequency ranges above beta, which may also carry relevant sleep-related information [114].

Finally, the sample size was modest, and the use of daily median values may have obscured transient fluctuations in beta activity that could be functionally relevant [106].

Despite these limitations, our study provides a longitudinal characterization of STN spectral dynamics across sleep and wake states under real-world conditions during both conventional and adaptive stimulation. By showing that sleep-related reductions in beta power are predominantly driven by aperiodic spectral components and remain independent of stimulation mode, our findings refine the interpretation of beta activity as a biomarker for aDBS and emphasize the importance of considering the full spectral architecture of STN signals. Incorporating aperiodic features into future adaptive algorithms may improve the physiological specificity of aDBS and support the development of circadian-informed, personalized neuromodulation strategies for PD. More broadly, these findings contribute to a deeper understanding of subthalamic physiology in naturalistic conditions and may help guide the next generation of adaptive DBS therapies.

## Data availability

The datasets presented in this article are not readily available because LFP recorded with the AlphaDBS device cannot be deposited in a public repository as they can be traceable to the identity of the subject. Requests to access the datasets should be directed to IUI.

## Code availability

No original method has been developed. All analyses were performed in MATLAB 2021a (The MathWorks Inc., Natik, MA, USA) with standard functions.

## Acknowledgements

The study was funded by the European Union - Next Generation EU - NRRP M6C2 - Investment 2.1 Enhancement and strengthening of biomedical research in the NHS, and by the Fondazione Pezzoli per la Malattia di Parkinson – ETS. RH, CP and IUI were supported by the Deutsche For-schungsgemeinschaft (DFG, German Research Foundation) Project-ID 424778381 - TRR 295.

FL, CP, PS, EC and IUI were supported by the Michael J. Fox Foundation for Parkinson’s Research.

FL, RH and LC were supported by a grant of the German Excellence Initiative to the Graduate School of Life Sciences, University of Würzburg.

AM was supported by the project “IMAD23ALM MAD: The etiopathological basis of gait derangement in Parkinson’s disease: decoding locomotor network dynamics”. AM acknowledges the support of the BRIEF “Biorobotics Research and Innovation Engineering Facilities” project (Project Identification Code IR0000036) funded under the National Recovery and Resilience Plan (NRRP), Mission 4 Component 2 Investment 3.1 of Italian Ministry of University and Research funded by the European Union–Next-Generation EU.

We thank Rebecca Barbiani and Ilaria Riela of the Fondazione Pezzoli for Parkinson’s Disease and the ASST G. Pini-CTO for their administrative support.

Newronika SpA. played no role in data analysis and interpretation, and the writing of this manuscript.

We thank Fabio Taddeini of the Biorobotics Institute of the Sant’Anna School of Advanced Studies for his valuable insights into data analysis.

AI-assisted language editing was used to improve grammar, clarity, and readability. The scientific content, interpretations, and conclusions were developed exclusively by the authors.

## Author contributions

Conceptualization: LC, FL, RH, CP, AM, IUI

Methodology: LC, CP, AM, IUI

Formal Analysis: LC, FL, CP, AM, IUI

Investigation: FL, PS, EC, SM, IUI

Data Curation: LC, FL, PS, EC, CP

Validation: LC, FL, CP

Resources: CP, AM, IUI

Writing: LC, CP, AM, IUI

Writing – Review & Editing: FL, SC, RH, SB, PS, EC, SM

Visualization: LC, AM

Supervision: CP, AM, IUI

Project Administration: IUI

Funding Acquisition: CP, AM, IUI

