## Supplementary Materials for "Aperiodic subthalamic activity underlies sleep-wake modulation of beta power during conventional and adaptive deep brain stimulation in Parkinson’s disease"

**Supplementary Figure 1: STN-LFP power spectral density with periodic and aperiodic component for all patients in active cDBS or aDBS.** Each panel represents a patient. Upper left: Loglog plot of median STN-LFP PSD (solid line) and median aperiodic component (dashed line) in cDBS, separately for sleep (brown) and wake (yellow) states. Shaded regions correspond to the interquartile range (first to third quartile). Upper right: Same as upper left but for aDBS. Lower left: Loglog plot of median STN-LFP periodic component (dotted line) in cDBS, separately for sleep (brown) and wake (yellow) states. Shaded regions correspond to the interquartile range (first to third quartile). Lower right: Same as lower left but for aDBS. Abbreviations: a, adaptive; c, conventional; DBS, deep brain stimulation; LFP, local field potentials; PSD, power spectral density; Pt., patient; STN, subthalamic nucleus.

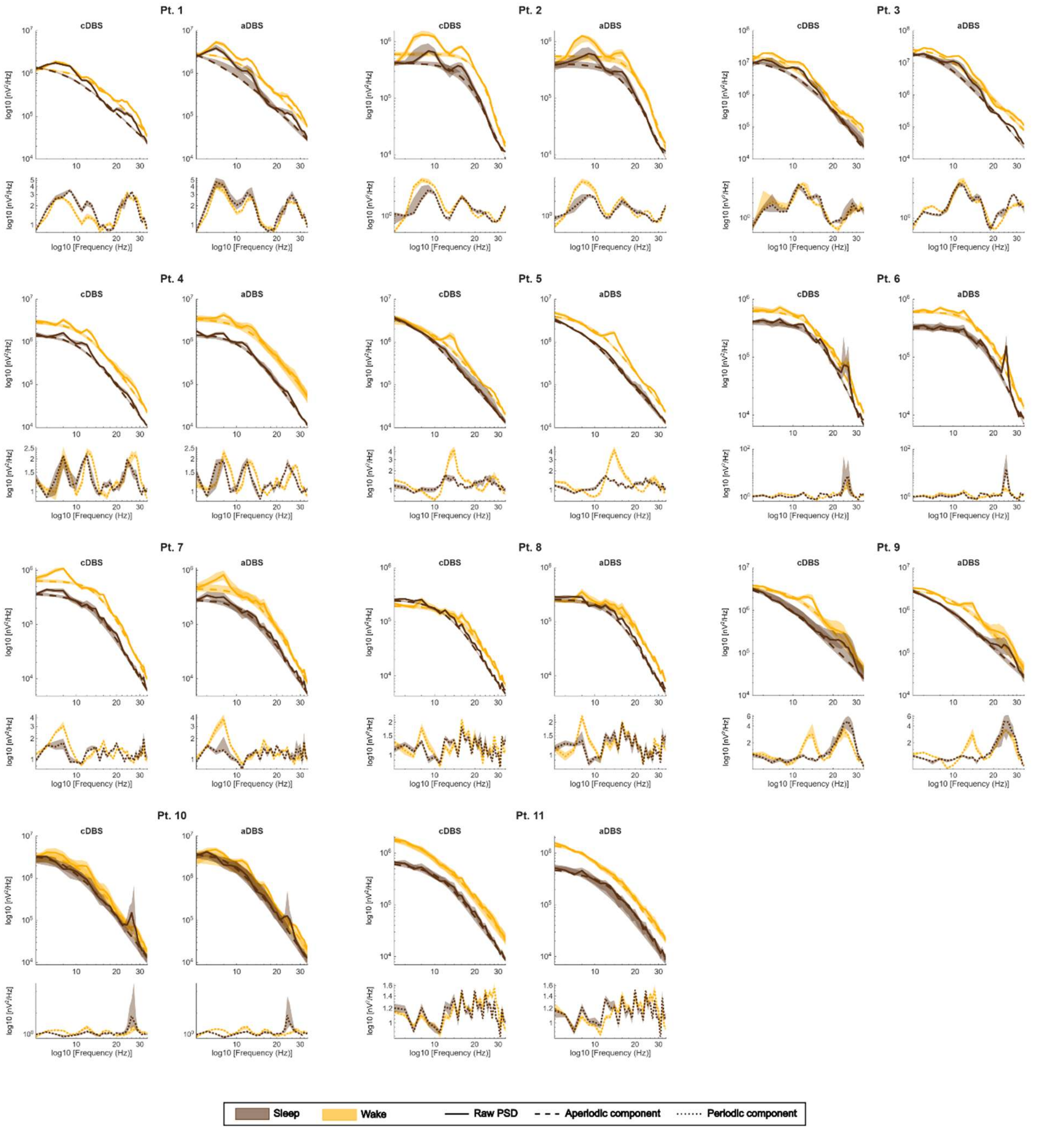

**Supplementary Figure 2: DBS leads localization.** 3D localization of the DBS electrodes for both left and right STN for all patients except pt. 2 and 11 for which the imaging were not available. The red nucleus is indicated in red color, while the STN in orange. DBS electrodes and contacts were localized based on pre- and postoperative neuroimaging using a tool designed for this task (as implemented in Lead-DBS software [1]). Abbreviations: DBS, deep brain stimulation; STN, subthalamic nucleus.

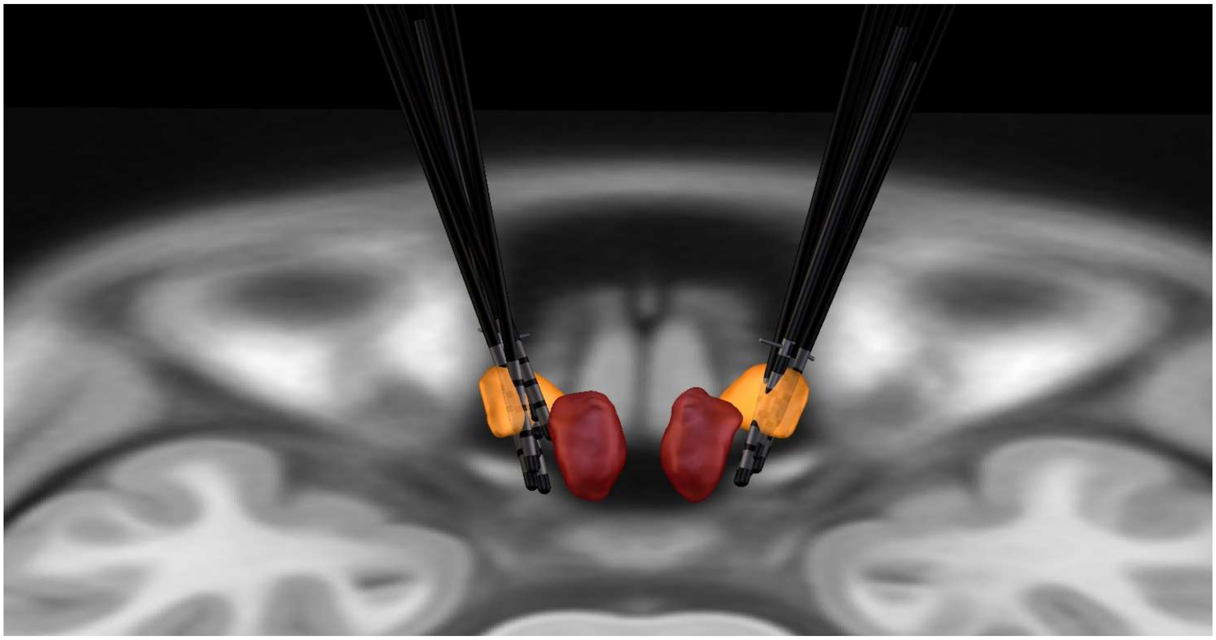

**Supplementary Figure 3: Time course of stimulation current in aDBS for patient 3.**
Stimulation current delivered every minute to the left and right STN is shown for each day of aDBS in patient 3, in the left and right panel, respectively. Grey boxes indicate time windows during which the current varied less than 0.2 mA for at least 60 consecutive minutes. Abbreviations: a, adaptive; DBS, deep brain stimulation; STN, subthalamic nucleus.

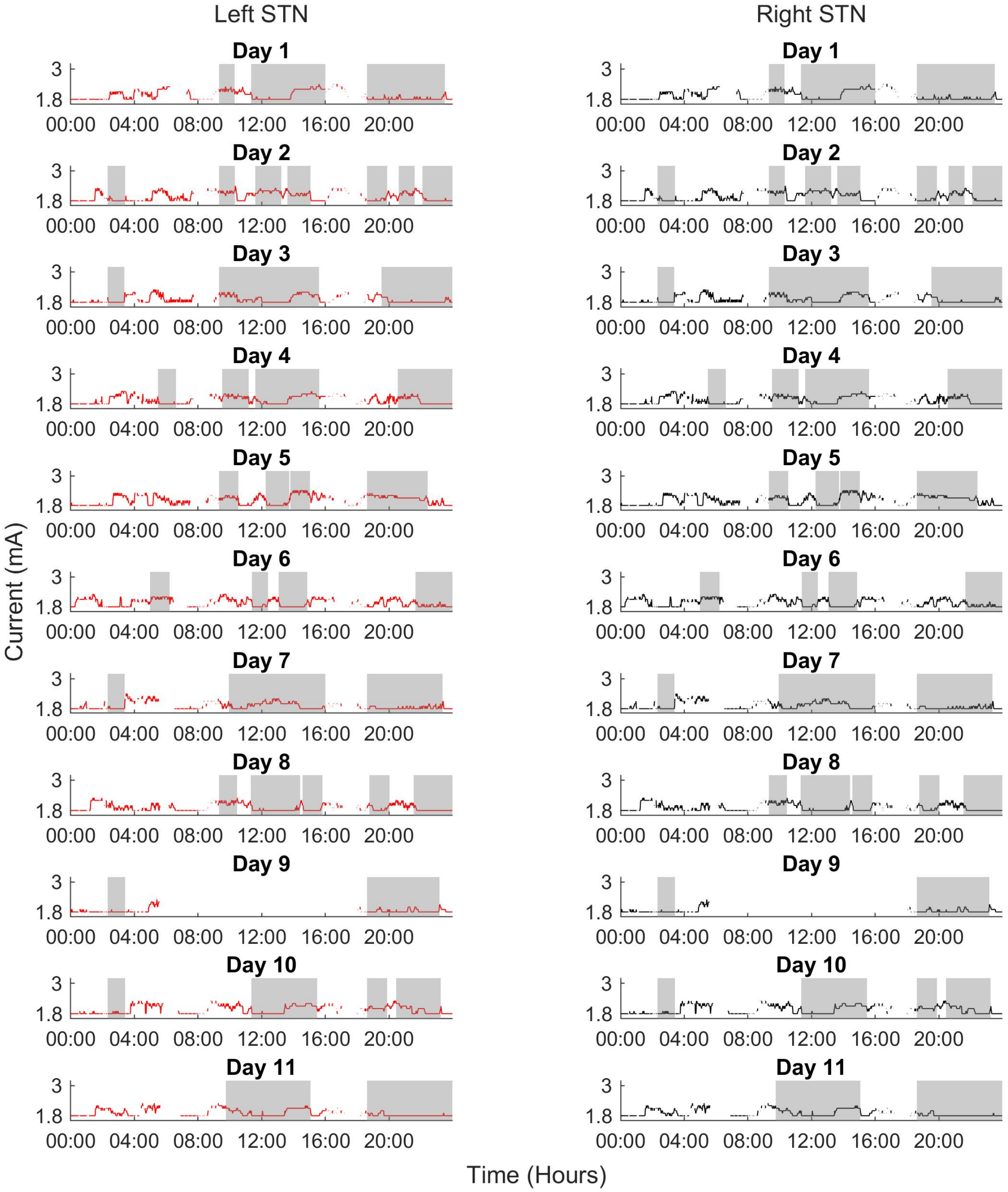

**Supplementary Figure 4: Effect of state and mode on each spectral feature (interquartile range).** For each spectral feature x, box chart of $x_{IQR}$ in cDBS and aDBS separately for the sleep (brown) and wake (yellow) state. Each dot represents the $x_{IQR}$ of an individual patient in the corresponding state and mode and grey dotted lines connect the same patient between sleep and wake state (light grey if wake > sleep and dark grey otherwise). The subtitle repots the results of the repeated measures ANOVA test with Benjamini & Hochberg false discovery rate correction (p>0.05: ns, 0.01<p<0.05: *, 0.001<p<0.01: **, and p<0.001: ***). Abbreviations: a, adaptive; $BP_{IQR}$, overall interquartile range of broadband power of the aperiodic component; c, conventional; DBS, deep brain stimulation; $KneeFreq_{IQR}$, overall interquartile range of knee frequency of the aperiodic component; $Offset_{IQR}$, overall interquartile range of offset of the aperiodic component; $pHBP_{IQR}$, overall interquartile range of power of periodic component in the high beta band; $pLBP_{IQR}$, overall interquartile range of power of periodic component in the low beta band; $pPP_{IQR}$, overall interquartile range of power of periodic component in the peak range; $rHBP_{IQR}$, overall interquartile range of power of raw PSD in the high beta band; $rLBP_{IQR}$, overall interquartile range of power of raw PSD in the low beta band; $rPP_{IQR}$, overall interquartile range of power of raw PSD in the peak range; $Slope_{IQR}$, overall interquartile range of slope of the aperiodic component.

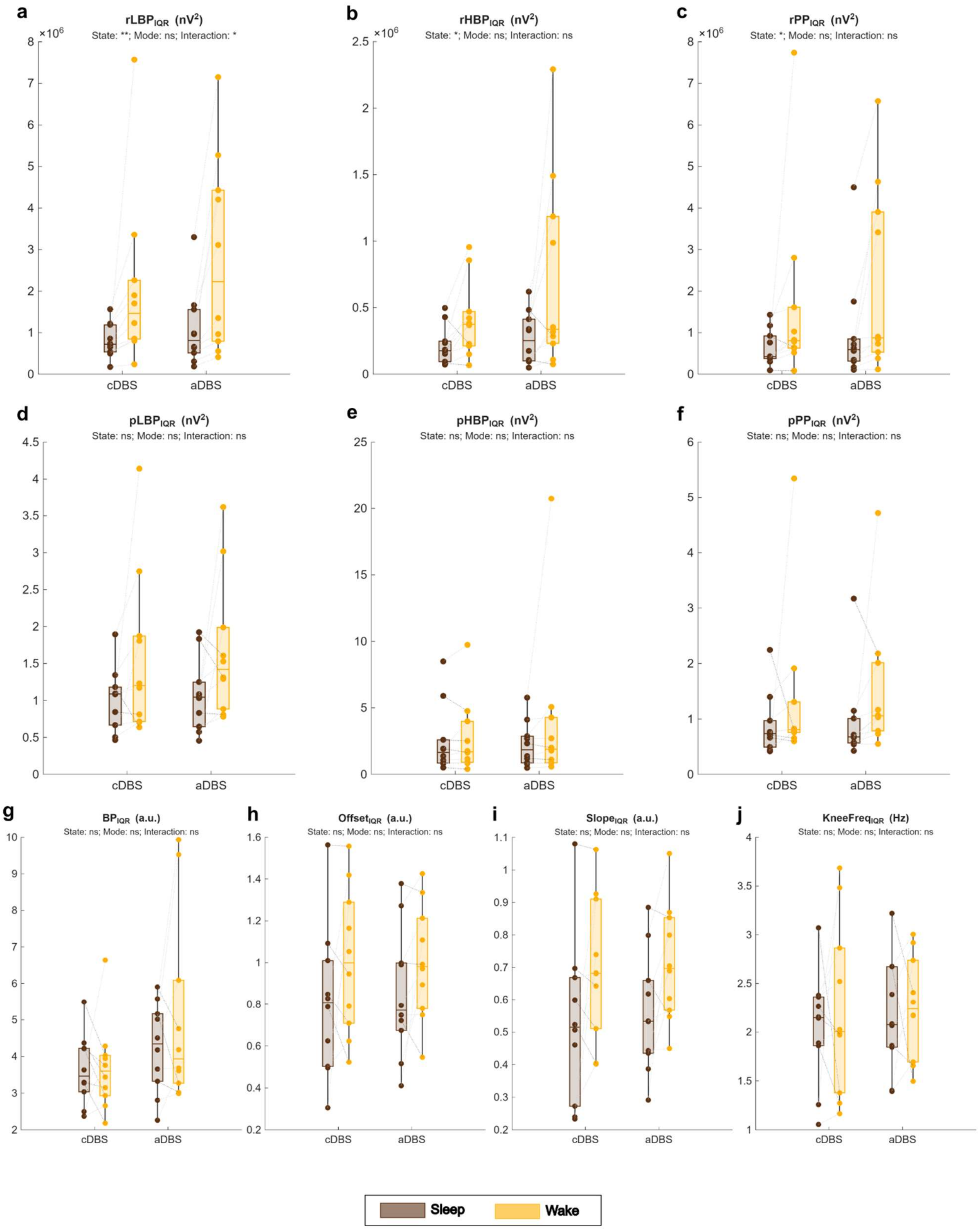
a
rLBP$_{IQR}$ (nV$^2$)
State: **; Mode: ns; Interaction: *
b
rHBP$_{IQR}$ (nV$^2$)
State: *; Mode: ns; Interaction: ns
c
rPP$_{IQR}$ (nV$^2$)
State: *; Mode: ns; Interaction: ns
d
pLBP$_{IQR}$ (nV$^2$)
State: ns; Mode: ns; Interaction: ns
e
pHBP$_{IQR}$ (nV$^2$)
State: ns; Mode: ns; Interaction: ns
f
pPP$_{IQR}$ (nV$^2$)
State: ns; Mode: ns; Interaction: ns
g
BP$_{IQR}$ (a.u.)
State: ns; Mode: ns; Interaction: ns
h
Offset$_{IQR}$ (a.u.)
State: ns; Mode: ns; Interaction: ns
i
Slope$_{IQR}$ (a.u.)
State: ns; Mode: ns; Interaction: ns
j
KneeFreq$_{IQR}$ (Hz)
State: ns; Mode: ns; Interaction: ns
cDBS
aDBS
Sleep
Wake

**Supplementary Table 1: Stimulation programming parameters in cDBS and aDBS.** Each row represents one patient. For each patient, the table reports from left to right: contacts selected for stimulation in the left (L) and right (R) STN; current amplitude used for cDBS in both hemispheres; stimulation frequency (F); stimulation pulse width (PW); current range for the adaptive stimulation, from minimum (Amin) to maximum (Amax) value in both hemispheres; contacts selected for recording in the STN that drives current adaptation in aDBS; beta amplitude range from minimum (βmin) to maximum (βmax) value used for linear current modulation in aDBS; frequency range monitored for aDBS; and median current and total electrical energy delivered (TEED) for the driver hemisphere in cDBS and in aDBS, separately for the sleep and wake states, as well as across the total recording time. Patients 1 to 8 were implanted with quadripolar 3389 lead (Medtronic), in which contact numbering follows the convention that contacts 0 and 8 are the ventralmost, and contacts 3 and 11 are the dorsalmost, for the left and right STN, respectively. Patients 9 to 11 were implanted with 6172 Directed leads (Abbott/St. Jude Medical), in which contact numbering follows the convention that contacts 1 and 9 are the ventralmost, and contacts 4 and 12 are the dorsalmost, for the left and right STN, respectively. The central contacts of the 6172 Directed leads are divided into 3 segments, labeled from a to c. In all patients, stimulation was delivered in a monopolar configuration, with the active contact serving as the cathode (-) and the implantable pulse generator case as the anode (+). Recordings were performed in a bipolar configuration, with the dorsalmost contact serving as the anode and the ventralmost contact as the cathode. Abbreviations: a, adaptive; A, current amplitude; β, average normalized beta amplitude; c, conventional; DBS, deep brain stimulation; F, frequency; L, left STN; Pt., patient; PW, pulse width; R, right STN; STN, subthalamic nucleus; TEED, total electrical energy delivered.

| | **Stimulating contacts** | | **AcDBS (mA)** | | **F (Hz)** | **PW (µs)** | **Amin – Amax (mA)** | | **Recording contacts (driver STN)** | **βmin – βmax (a.u.)** | **F range (Hz)** | **A (mA) – TEED (µW)** | | | |
|---|---|---|---|---|---|---|---|---|---|---|---|---|---|---|---|
| | **L** | **R** | **L** | **R** | | | **L** | **R** | | | | **cDBS** | **aDBS Sleep** | **aDBS Wake** | **aDBS Total** |
| Pt. 1 | 1 | 8 | 2.7 | 2.8 | 130 | 80 | 2.2 – 3.1 | 2.3 – 2.8 | 0 – 2 (L) | 215 – 250 | 11 – 16 | 2.7 – 75.8 | 2.6 – 70.3 | 3.0 – 93.6 | 2.9 – 87.5 |
| Pt. 2 | 2 | 8 | 4.0 | 3.5 | 70 | 60 | 3.5 – 4.5 | 3.0 – 4.0 | 9 – 10 (R) | 330 – 355 | 12 – 19 | 3.5 – 51.5 | 3.1 – 40.4 | 3.6 – 54.4 | 3.4 – 48.6 |
| Pt. 3 | 2 | 9 | 2.2 | 2.1 | 130 | 60 | 1.8 – 3.0 | 1.8 – 3.0 | 8 – 10 (R) | 240 – 350 | 12 – 18 | 2.1 – 34.4 | 1.8 – 25.3 | 1.9 – 28.2 | 1.8 – 25.3 |
| Pt. 4 | 0 | 10 | 4.0 | 4.0 | 80 | 60 | 2.5 – 4.5 | 3.0 – 4.5 | 1 – 2 (L) | 280 – 310 | 11 – 17 | 4.0 – 76.8 | 3.7 – 65.7 | 3.8 – 69.3 | 3.8 – 69.3 |
| Pt. 5 | 2 | 9 | 2.4 | 2.3 | 130 | 60 | 2.0 – 2.9 | 2.0 – 3.5 | 1 – 3 (L) | 295 – 425 | 11 – 20 | 2.4 – 44.9 | 2.1 – 34.4 | 2.5 – 48.8 | 2.4 – 44.9 |
| Pt. 6 | 3 | 11 | 2.0 | 2.2 | 130 | 60 | 1.5 – 2.6 | 1.5 – 2.6 | 9 – 10 (R) | 335 – 405 | 9 – 16 | 2.2 – 37.8 | 1.9 – 28.2 | 2.1 – 34.4 | 2.1 – 34.4 |
| Pt. 7 | 3 | 10 | 2.3 | 1.8 | 130 | 60 | 1.5 – 3.0 | 1.2 – 1.8 | 1 – 2 (L) | 310 – 360 | 12 – 20 | 2.3 – 41.3 | 2.1 – 34.4 | 2.4 – 44.9 | 2.2 – 37.8 |
| Pt. 8 | 2 | 9 | 2.4 | 3.6 | 180 | 60 | 2.0 – 2.6 | 3.0 – 4.0 | 10 – 11 (R) | 130 – 180 | 19 – 25 | 3.6 – 140.0 | 3.3 – 117.6 | 3.7 – 147.9 | 3.7 – 147.9 |
| Pt. 9 | 2b | 11b | 1.4 | 1.4 | 130 | 60 | 0.8 – 2.0 | 1.0 – 2.5 | 2a – 3a (L) | 210 – 430 | 12 – 18 | 1.4 – 15.3 | 0.8 – 5.0 | 1.1 – 9.4 | 0.9 – 6.3 |
| Pt. 10 | 3c | 11b | 1.8 | 2.4 | 130 | 60 | 1.7 – 2.5 | 2.0 – 3.0 | 9 – 12 (R) | 225 – 270 | 12 – 18 | 2.4 – 44.9 | 2.3 – 41.3 | 2.6 – 52.7 | 2.6 – 52.7 |
| Pt. 11 | 3c | 12 | 2.2 | 2.2 | 130 | 60 | 2.0 – 2.5 | 2.0 – 2.5 | 3a – 3b (L) | 200 – 250 | 12 – 17 | 2.2 – 37.8 | 2.4 – 44.9 | 2.3 – 41.3 | 2.2 – 37.8 |

**Supplementary Table 2: Current amplitude modulation in aDBS mode**. Each row represents one patient. For each patient, the table reports the average daily occurrence (expressed as a percentage of the sleep, wake, or total time) and the duration (in minutes) of time windows during which the stimulation current of the driver STN (i.e., the one controlling current adaptation in aDBS) varied less than 0.2 mA for at least 60 consecutive minutes, separately for the wake and sleep states, as well as for the total time. Abbreviations: a, adaptive; DBS, deep brain stimulation; min, minutes; Pt., patient; STN, subthalamic nucleus.

| | **Occurrence (%)** | | | **Duration (min)** | | |
|---|---|---|---|---|---|---|
| | **Sleep** | **Wake** | **Total** | **Sleep** | **Wake** | **Total** |
| Pt.1 | 8.3 | 2.9 | 4.0 | 75.8 | 72.7 | 78.2 |
| Pt. 2 | 16.8 | 2.0 | 6.4 | 91.2 | 78.5 | 90.2 |
| Pt. 3 | 58.9 | 76.8 | 73.7 | 109.6 | 179.0 | 181.0 |
| Pt. 4 | 0.0 | 0.0 | 0.6 | – | – | 82.0 |
| Pt. 5 | 41.6 | 3.3 | 14.8 | 95.4 | 67.8 | 78.4 |
| Pt. 6 | 0.0 | 0.0 | 0.8 | – | – | 105.0 |
| Pt. 7 | 0.0 | 0.0 | 0.0 | – | – | – |
| Pt. 8 | 2.8 | 1.0 | 1.2 | 67.0 | 61.0 | 64.0 |
| Pt. 9 | 94.5 | 14.8 | 40.2 | 206.9 | 83.0 | 155.3 |
| Pt. 10 | 35.5 | 0.8 | 9.7 | 130.1 | 62.0 | 133.2 |
| Pt. 11 | 54.4 | 31.4 | 37.7 | 164.2 | 88.1 | 104.8 |

**Supplementary Table 3: Hauser diary.** Each row represents one patient. For each patient, the table reports, separately for cDBS and aDBS, the average time in hours spent in sleep state, OFF state (OFF), ON state without any dyskinesia ($ON_{NoDysk}$), ON state without troublesome dyskinesia (good-on-time, GOT; defined as ON time without dyskinesia plus ON time with mild dyskinesia), and ON state with dyskinesia ($ON_{NoDysk}$; defined as the sum of ON time with mild, moderate and severe dyskinesia). Values were calculated as the average across the three days during which patients completed the Hauser diary in each stimulation mode. Percentage values relative to wake time were obtained by dividing the time spent in each state by the wake time and multiplying by 100. Wake time was calculated as 24 hours minus total sleep time. Abbreviations: a, adaptive; c, conventional; DBS, deep brain stimulation; GOT, good-on-time; h, hours; OFF, OFF state; $ON_{Dysk}$, ON state with dyskinesia; $ON_{NoDysk}$, ON state without dyskinesia, Pt., patient.

| | **cDBS** | | | | | **aDBS** | | | | |
|---|---|---|---|---|---|---|---|---|---|---|
| | **Sleep (h)** | **OFF (h)** | **$ON_{NoDysk}$ (h)** | **GOT (h)** | **$ON_{Dysk}$ (h)** | **Sleep (h)** | **OFF (h)** | **$ON_{NoDysk}$ (h)** | **GOT (h)** | **$ON_{Dysk}$ (h)** |
| Pt.1 | 7.5 | 7.7 | 8.2 | 8.8 | 0.0 | 8.3 | 0.0 | 15.7 | 15.7 | 0.0 |
| Pt. 2 | 6.5 | 3.0 | 10.3 | 14.0 | 0.5 | 9.2 | 3.8 | 7.8 | 11.0 | 0.0 |
| Pt. 4 | 8.8 | 4.2 | 10.2 | 11.0 | 0.0 | 6.8 | 6.0 | 11.2 | 11.2 | 0.0 |
| Pt. 5 | 7.3 | 7.5 | 8.8 | 9.2 | 0.0 | 7.7 | 0.0 | 15.7 | 16.3 | 0.0 |
| Pt. 6 | 5.7 | 14.7 | 3.7 | 3.7 | 0.0 | 5.5 | 0.0 | 18.5 | 18.5 | 0.0 |
| Pt. 7 | 9.2 | 6.8 | 8.0 | 8.0 | 0.0 | 10.7 | 9.3 | 4.0 | 4.0 | 0.0 |
| Pt. 8 | 8.3 | 0.0 | 11.3 | 15.2 | 0.5 | 7.5 | 0.0 | 13.7 | 16.3 | 0.2 |
| Pt. 9 | 5.7 | 2.0 | 16.3 | 16.3 | 0.0 | 5.7 | 0.0 | 17.7 | 18.3 | 0.0 |
| Pt. 10 | 6.7 | 5.5 | 0.2 | 9.7 | 0.0 | 7.2 | 0.0 | 1.8 | 16.8 | 0.0 |
| Pt. 11 | 7.0 | 1.7 | 10.8 | 15.3 | 0.0 | 7.7 | 0.8 | 13.7 | 15.5 | 0.0 |
| **Mean** | **7.3** | **5.3** | **8.8** | **11.1** | **0.1** | **7.6** | **2** | **12** | **14.4** | **0** |
| **[min – max]** | **[5.7 – 9.2]** | **[0 – 14.7]** | **[0.2 – 16.3]** | **[3.7 – 16.3]** | **[0 – 0.5]** | **[5.5 – 10.7]** | **[0 – 9.3]** | **[1.8 – 18.5]** | **[4 – 18.5]** | **[0 – 0.2]** |

**Supplementary Table 4: Time spent in wake and sleep states.** Each row represents one patient. For each patient, the table reports the average daily sleep and wake time. Wake-up and bedtime information was derived from patients' diaries using a conservative approach, ensuring that the selected time intervals corresponded unambiguously to wake or sleep state. Abbreviations: Pt., patient.

| | **Monitoring Time<br>Start time – End time (Total time in hours)** | |
|---|---|---|
| | **Sleep** | **Wake** |
| Pt.1 | 00:00 – 06:00 (6) | 10:00 – 22:00 (12) |
| Pt. 2 | 00:00 – 06:00 (6) | 09:30 – 22:00 (12.5) |
| Pt. 4 | 22:00 – 05:00 (7) | 07:00 – 20:00 (13) |
| Pt. 5 | 23:30 – 04:00 (4.5) | 07:00 – 21:00 (14) |
| Pt. 6 | 01:00 – 04:00 (3) | 08:00 – 22:00 (14) |
| Pt. 7 | 23:00 – 05:30 (6.5) | 10:00 – 20:30 (10.5) |
| Pt. 8 | 02:00 – 07:00 (5) | 10:00 – 24:00 (14) |
| Pt. 9 | 01:00 – 05:00 (4) | 07:00 – 23:00 (16) |
| Pt. 10 | 01:00 – 05:30 (4.5) | 07:30 – 23:00 (15.5) |
| Pt. 11 | 01:00 – 06:30 (5.5) | 09:00 – 22:00 (13) |
| **Mean [min – max]** | **5.2 [3 – 7]** | **13.5 [10.5 – 16]** |

**Supplementary Table 5: Statistical comparison of $x_{MED}$.** Each row represents a spectral feature. For each spectral features, the table reports the results of the two-factor, within-subject repeated measures ANOVA. Effect sizes are reported in brackets in case of statistical significance (partial eta squared). See the *Methods* section for further details. Abbreviations: a, adaptive; $BP_{MED}$, overall median broadband power of the aperiodic component; c, conventional; DBS, deep brain stimulation; $KneeFreq_{MED}$, overall median knee frequency of the aperiodic component; $Offset_{MED}$, overall median offset of the aperiodic component; $pHBP_{MED}$, overall median power of periodic component in the high beta band; $pLBP_{MED}$, overall median power of periodic component in the low beta band; $pPP_{MED}$, overall median power of periodic component in the peak range; $rHBP_{MED}$, overall median power of raw PSD in the high beta band; $rLBP_{MED}$, overall median power of raw PSD in the low beta band; $rPP_{MED}$, overall median power of raw PSD in the peak range; $Slope_{MED}$, overall median slope of the aperiodic component.

| | **Repeated measures ANOVA** | | |
|---|---|---|---|
| | **State** | **Mode** | **Interaction** |
| $rLBP_{MED}$ | 0.0024 (0.69) | 0.40 | 0.50 |
| $rHBP_{MED}$ | 0.0015 (0.76) | 0.40 | 0.34 |
| $rPP_{MED}$ | 0.0033 (0.64) | 0.40 | 0.49 |
| $pLBP_{MED}$ | 0.14 | 0.95 | 0.53 |
| $pHBP_{MED}$ | 1.00 | 0.054 (0.48) | 0.53 |
| $pPP_{MED}$ | 0.14 | 0.70 | 0.53 |
| $BP_{MED}$ | < 0.001 (0.90) | 0.73 | 0.63 |
| $Offset_{MED}$ | 0.031 (0.49) | 0.73 | 0.81 |
| $Slope_{MED}$ | 0.16 | 0.73 | 1.00 |
| $KneeFreq_{MED}$ | 0.031 (0.45) | 0.92 | 0.81 |

**Supplementary Table 6: Statistical comparison of $x_{IQR}$.** Each row represents a spectral feature. For each spectral features, the table reports the results of the two-factor, within-subject repeated measures ANOVA. Effect sizes are reported in brackets in case of statistical significance (partial eta squared). See the *Methods* section for further details. Abbreviations: a, adaptive; $BP_{IQR}$, overall interquartile range of broadband power of the aperiodic component; c, conventional; DBS, deep brain stimulation; $KneeFreq_{IQR}$, overall interquartile range of knee frequency of the aperiodic component; $Offset_{IQR}$, overall interquartile range of offset of the aperiodic component; $pHBP_{IQR}$, overall interquartile range of power of periodic component in the high beta band; $pLBP_{IQR}$, overall interquartile range of power of periodic component in the low beta band; $pPP_{IQR}$, overall interquartile range of power of periodic component in the peak range; $rHBP_{IQR}$, overall interquartile range of power of raw PSD in the high beta band; $rLBP_{IQR}$, overall interquartile range of power of raw PSD in the low beta band; $rPP_{IQR}$, overall interquartile range of power of raw PSD in the peak range; $Slope_{IQR}$, overall interquartile range of slope of the aperiodic component.

| | Repeated measures ANOVA | | |
|---|---|---|---|
| | **State** | **Mode** | **Interaction** |
| $rLBP_{IQR}$ | 0.002 (0.75) | 0.19 | 0.019 (0.58) |
| $rHBP_{IQR}$ | 0.014 (0.50) | 0.11 | 0.10 |
| $rPP_{IQR}$ | 0.014 (0.52) | 0.25 | 0.092 |
| $pLBP_{IQR}$ | 0.10 | 0.43 | 0.31 |
| $pHBP_{IQR}$ | 0.32 | 0.43 | 0.40 |
| $pPP_{IQR}$ | 0.10 | 0.43 | 0.30 |
| $BP_{IQR}$ | 0.77 | 0.50 | 0.84 |
| $Offset_{IQR}$ | 0.23 | 0.77 | 0.84 |
| $Slope_{IQR}$ | 0.18 | 0.77 | 0.84 |
| $KneeFreq_{IQR}$ | 0.77 | 0.77 | 0.84 |
